# Multi-Ancestry Transcriptome-wide Association Studies Uncover New Insights into Breast Cancer Genetics and Biology

**DOI:** 10.1101/2025.08.12.25333509

**Authors:** Jie Ping, Guochong Jia, Qiuyin Cai, Xingyi Guo, Jifeng Wang, Ran Tao, Bingshan Li, Joshua A. Bauer, Yuhan Xie, Stefan Ambs, Mollie E. Barnard, Yu Chen, Ji-Yeob Choi, Yu-Tang Gao, Montserrat Garcia-Closas, Jian Gu, Jennifer J. Hu, Motoki Iwasaki, Esther M. John, Sun-Seog Kweon, Christopher I. Li, Koichi Matsuda, Keitaro Matsuo, Katherine L. Nathanson, Barbara Nemesure, Olufunmilayo I. Olopade, Tuya Pal, Sue K. Park, Boyoung Park, Michael F. Press, Maureen Sanderson, Dale P. Sandler, Song Yao, Ying Zheng, Thomas Ahearn, Abenaa M. Brewster, Adeyinka Falusi, Anselm J.M. Hennis, Hidemi Ito, Michiaki Kubo, Eun-Sook Lee, Timothy Makumbi, Berthe S.E. Mapoko, Dong-Young Noh, Katie M. O’Brien, Oladosu Ojengbede, Andrew F. Olshan, Min-Ho Park, Sonya Reid, Taiki Yamaji, Gary Zirpoli, Ebonee N. Butler, Maosheng Huang, Siew-Kee Low, John Obafunwa, Clarice R. Weinberg, Haoyu Zhang, Hongyu Zhao, Christine B. Ambrosone, Michelle L. Cote, Dezheng Huo, Christopher A. Haiman, Daehee Kang, Julie R. Palmer, Melissa A. Troester, Xiao-Ou Shu, Jirong Long, Wei Zheng

**Affiliations:** Division of Epidemiology, Department of Medicine, Vanderbilt Epidemiology Center, Vanderbilt-Ingram Cancer Center, Vanderbilt University Medical Center, Nashville, TN, USA; Department of Public Health Sciences, University of Miami School of Medicine, Miami, FL, USA; Department of Biostatistics, Vanderbilt University Medical Center, Nashville, TN, USA; Department of Molecular Physiology & Biophysics, Vanderbilt Genetics Institute, Vanderbilt University, Nashville, TN, USA; Department of Biochemistry, Vanderbilt Institute of Chemical Biology, Vanderbilt University School of Medicine, Nashville, TN, USA; Department of Biostatistics, Yale School of Public Health, New Haven, CT, USA; Laboratory of Human Carcinogenesis, Center of Cancer Research, National Cancer Institute, National Institutes of Health, Bethesda, MD, USA; Slone Epidemiology Center, Boston University, Boston, MA, USA; Division of Epidemiology, Department of Population Health, NYU Grossman School of Medicine, New York, NY, USA; Department of Biomedical Sciences, Seoul National University College of Medicine, Seoul, Republic of Korea; Department of Preventive Medicine, Seoul National University College of Medicine, Seoul, Republic of Korea; State Key Laboratory of Oncogene and Related Genes & Department of Epidemiology, Shanghai Cancer Institute, Renji Hospital, Shanghai Jiao Tong University School of Medicine, Shanghai, China; Division of Cancer Epidemiology and Genetics, National Cancer Institute, Bethesda, MD, USA; Department of Epidemiology, The University of Texas MD Anderson Cancer Center, Houston, TX, USA; Division of Epidemiology, National Cancer Center Institute for Cancer Control, Tokyo, Japan; Departments of Epidemiology & Population Health and of Medicine, Stanford University School of Medicine, Stanford, CA, USA; Department of Preventive Medicine, Chonnam National University Medical School, Hwasun, Republic of Korea; Jeonnam Regional Cancer Center, Chonnam National University Hwasun Hospital, Hwasun, Republic of Korea; Division of Public Health Sciences, Fred Hutchinson Cancer Center, Seattle, WA, USA; Laboratory of Clinical Genome Sequencing, Graduate School of Frontier Sciences, The University of Tokyo, Tokyo, Japan; Division of Cancer Epidemiology and Prevention, Aichi Cancer Center Research Institute, Nagoya, Japan; Division of Cancer Epidemiology, Nagoya University Graduate School of Medicine, Nagoya, Japan; Division of Hematology/Oncology, Department of Medicine, Perelman School of Medicine, University of Pennsylvania, Philadelphia, PA, USA; Basser Center for BRCA, Abramson Cancer Center, Perelman School of Medicine, University of Pennsylvania, Philadelphia, PA, USA; Department of Family, Population and Preventive Medicine, Renaissance School of Medicine, Stony Brook University, Stony Brook, NY, USA; Center for Clinical Cancer Genetics and Global Health, Department of Medicine, The University of Chicago, Chicago, IL, USA; Division of Genetic Medicine, Department of Medicine, Vanderbilt-Ingram Cancer Center, Vanderbilt University Medical Center, Nashville, TN, USA; Integrated Major in Innovative Medical Science, Seoul National University Graduate School, Jongnogu, Seoul, Republic of Korea; Cancer Research Institute, Seoul National University, Jongnogu, Seoul, Republic of Korea; Department of Preventive Medicine, Hanyang University College of Medicine, Seoul, Republic of Korea; Department of Pathology, Norris Comprehensive Cancer Center, University of Southern California, Los Angeles, CA, USA; Department of Family and Community Medicine, Meharry Medical College, Nashville, TN, USA; Epidemiology Branch, National Institute of Environmental Health Sciences, National Institutes of Health, Research Triangle Park, NC, USA; Department of Cancer Prevention and Control, Roswell Park Comprehensive Cancer Center, Elm & Carlton Streets, Buffalo, NY, USA; Shanghai Municipal Center for Disease Control and Prevention, Shanghai, China; Department of Clinical Cancer Prevention, The University of Texas MD Anderson Cancer Center, Houston, TX, USA; Genetic and Bioethics Research Unit, Institute for Advanced Medical Research and Training, College of Medicine, University of Ibadan, Ibadan, Nigeria; George Alleyne Chronic Disease Research Centre, University of the West Indies, Bridgetown, Barbados; Division of Cancer Information and Control, Aichi Cancer Center Research Institute, Nagoya, Japan; Department of Descriptive Cancer Epidemiology, Nagoya University Graduate School of Medicine, Nagoya, Japan; RIKEN Center for Integrative Medical Sciences, Yokohama, Japan; National Cancer Center Graduate School of Cancer Science and Policy, Goyang, Republic of Korea; Hospital, National Cancer Center, Goyang, Republic of Korea; Department of Surgery, Mulago Hospital, Kampala, Uganda; Yaounde Central Hospital, Yaounde, Cameroon; College of Medicine, Cancer Research Institute, Seoul National University, Seoul, Republic of Korea; Department of Surgery, Seoul National University Hospital, Seoul, Republic of Korea; Center for Population and Reproductive Health, College of Medicine, University of Ibadan, Ibadan, Nigeria; Department of Epidemiology and Lineberger Comprehensive Cancer Center, University of North Carolina at Chapel Hill, Chapel Hill, NC, USA; Department of Surgery, Chonnam National University Medical School, Republic of Korea; Division of Hematology and Oncology, Department of Medicine, Vanderbilt-Ingram Cancer Center, Vanderbilt University Medical Center, Nashville, TN, USA; Department of Pathology and Forensic Medicine, Lagos State University Teaching Hospital, Lagos, Nigeria; Biostatistics and Computational Biology Branch, National Institutes of Environmental Health Sciences, Research Triangle Park, NC, USA; Fairbanks School of Public Health, Indiana University, Indianapolis, IN, USA; Simon Comprehensive Cancer Center, Indianapolis, IN, USA; Department of Public Health Sciences, The University of Chicago, Chicago, IL, USA; Department of Preventive Medicine, Keck School of Medicine of USC, Los Angeles, CA, USA

**Author notes:** **Corresponding author contact information:** Wei Zheng, M.D., Ph.D. Vanderbilt Epidemiology Center, Vanderbilt University School of Medicine, 2525 West End Avenue, 8th Floor, Nashville, TN 37203-1738.

**Keywords:** breast cancer, TWAS, susceptibility gene, multi-ancestry

## Abstract

Genome-wide association studies (GWAS) have identified over 200 genetic risk loci for breast cancer, yet the target genes in these loci remain largely unknown. To address this knowledge gap, we conducted a series of multi-ancestry transcriptome-wide association studies (TWAS) to discover potential breast cancer susceptibility genes. We developed and validated ancestry-specific genetic models to predict levels of gene expression, alternative splicing, and 3’ UTR alternative polyadenylation, using genomic and transcriptomic data from normal breast tissue samples of 652 females of African, Asian, or European ancestry. These models were then applied to GWAS data of 178,534 breast cancer cases and 248,300 controls from these ancestry groups for association analyses. We identified 290 genes associated with breast cancer risk, including 103 previously unreported in TWAS and 46 located at least 500Kb away from any previously identified risk variants. Among them, 39 genes exhibited distinct associations with breast cancer risk by estrogen receptor status. The identified genes were enriched in pathways related to homologous recombination, apoptosis, p53, PI3K/AKT/mTOR, estrogen, and IL-2/STAT5 signaling. Single-cell RNA sequencing and *in vitro* experiment data provided additional functional evidence for 169 genes. Our study uncovered large numbers of candidate breast cancer susceptibility genes and contributed valuable insights into the genetics and biology of this common cancer.

## Introduction

Breast cancer is the most common cancer among women globally,^1^ and genetic factors play a significant role in breast cancer etiology.^2^ Over the past two decades, genome-wide association studies (GWAS) have identified more than 200 genetic variants associated with breast cancer risk.^3–11^ However, these variants explain only 18% of familial relative risk and most of them are located in non-coding regions, making GWAS results difficult to interpret.^9,12^ By integrating genomics and transcriptomic data, transcriptome-wide association studies (TWAS) directly investigate gene-trait associations, providing added insights into the genetics of the phenotype of interest.^13,14^ Since 2017, we have conducted several TWAS of breast cancer.^2,15–20^ These studies, along with those conducted by others,^21–29^ have reported large numbers of associations between predicted gene expression and breast cancer risk. However, because limited transcriptomic data from normal breast tissue samples were available, virtually all previous TWAS primarily used transcriptomic data from other tissues in building gene prediction models, and thus the relevance of previous findings to breast cancer is a significant concern, particularly for genes not expressed in breast tissues. Furthermore, with a few exceptions,^15,16,30^ most TWAS of breast cancer were conducted among women of European ancestry.^2,17–29^ We have shown recently that TWAS conducted in non-European women could uncover novel genetic factors that are difficult to identify in studies of European-ancestry women.^15,16^ By combining genomic and transcriptomic data from all three ancestry groups, we expect to identify additional association signals and provide data for causal inferences.

Most TWAS of breast cancer conducted to date focused on investigating genetically regulated gene expression in relation to risk of this common cancer. Understanding that gene regulation involves more than just expression levels, we expanded our TWAS to include the analysis of post-transcriptional modifications, such as alternative splicing (AS) and alternative polyadenylation (APA), which are believed to play key roles in influencing gene function and phenotypes.^31,32^ Splicing TWAS (spTWAS) and APA-wide association studies (APA-WAS) investigate association of genetically regulated AS and APA, respectively, and capture post-transcriptional events that cannot be interrogated in traditional expression-based TWASs (Exp-TWAS). Herein, we present findings from a large multi-ancestry TWAS to identify putative breast cancer susceptibility genes by systematically evaluating associations of breast cancer risk with gene expression, AS and APA across the transcriptome.

## Results

We generated transcriptomic data using normal breast tissue samples from 156 women included in the Asia Breast Cancer Consortium (ABCC) and 381 women included in the Susan G. Komen Normal Tissue Bank (Komen). These data, along with genomic data from these women, were combined with data from the Genotype-Tissue Expression (GTEx) project (n=115) to build ancestry-specific models to predict gene expression, AS, and APA. Included in the model building were 150, 205, and 297 women of African, Asian, and European ancestry, respectively. For association analyses, we used data from GWAS of breast cancer conducted in 178,534 cases and 248,300 controls,^33^ including 18,034 cases and 22,104 controls of African descent from the African-ancestry Breast Cancer Genetic (AABCG), 27,116 cases and 112,407 controls of Asian descent from the ABCC and 133,384 cases and 113,789 controls of European descent from the Breast Cancer Association Consortium (BCAC). A detailed description of these data is provided in **Table S1**.

The overall study design is depicted in **Figure S1**. Since genetic architectures vary considerably across ancestry groups, we built ancestry-specific prediction models to predict gene expression by ancestry. For European ancestry, we used data from Komen (n=182) and GTEx (n=115); for Asian ancestry, we used data from Komen (n=49) and ABCC (n=156); and for African ancestry, we used data from Komen (n=150). In addition to building breast tissue-specific models using elastic net (⍰=0.5) using normal breast tissue transcriptome data from each source, we also used the Joint-Tissue Imputation (JTI) method to leverage transcriptome data from 33 tissue types from the GTEx to build a set of cross-tissue models.^34^ To improve the stability in estimating model parameters, we also built cross-study models for European and Asian descents (European: Komen-GTEx; Asian: ABCC-Komen) by using a modified version of the UTMOST (Unified Test for MOlecular SignaTures) framework to integrate data from two distinct datasets.^35,36^ We evaluated the validity of prediction models by comparing the predicted gene expression using models derived from a dataset with the observed gene expression from another dataset. For example, we predicted the expression level of each gene in GTEx European-ancestry dataset using the model derived from Komen European-ancestry dataset for that gene and compared the observed expression level of that gene in GTEx European-ancestry dataset to estimate a R^2^. We performed this correlation analysis for all 8,926 genes that can be predicted with a R^2^ of 0.01 or higher in both datasets and showed a correlation coefficient of 0.87, supporting the validity of the models built for the study (**Figure S2**).

Because multiple approaches were used to build the prediction models, it is possible that two or more models were built successfully to predict each of the three transcriptome events (gene expression, AS and APA) with an R^2^≥0.01. When this occurred, models with the highest prediction R^2^ were selected for downstream association analyses within each ancestry population. This led to the inclusion of 16,310 models for gene expression, 44,529 models for predicting AS in 11,964 genes, and 9,112 models for predicting APA in 6,457 genes for European-ancestry participants (Table S1). Similarly, we included 11,075 gene expression models, 29,591 models for predicting AS in 9,518 genes, and 7,688 models for predicting APA in 5,526 genes for Asian-ancestry participants, and included 8,452 gene expression models, 19,055 models for predicting AS in 7,970 genes, and 4,204 models for predicting APA in 3,229 genes for African-ancestry participants. Altogether, prediction models were built for 21,195 gene expression, 70,541 AS in 13,161 genes, and 14,730 APA in 9,499 genes, for at least one ancestral group.

We applied the prediction models built for each ancestry group to their corresponding GWAS data to perform association analyses using S-PrediXcan and then combined data using inverse-variance weighting meta-analyses. Of the 21,195 genes that can be predicted using our expression models, 11,087 genes were predicted in at least two ancestry groups, with 68 genes associated with overall breast cancer risk at a Bonferroni-corrected significance level of *2.4×10^−6^* (0.05/21,195) (**Figure 1A**, **Table S2**). Moreover, we identified 56 AS in 44 genes and 22 APA in 21 genes showing associations with overall breast cancer risk at Bonferroni-corrected significance levels of *7.1×10^−7^* (0.05/70,541 ASs, **Figure 1B**, **Table S3**) and *3.4×10^−6^* (0.05/14,730 APA, **Figure 1C**, **Table S4**). In ancestry-specific analyses, we identified an additional 80 genes, 114 AS in 87 genes, and 34 APA in 30 genes showing a significant association with risk of overall breast cancer at Bonferroni-corrected significance levels of *2.4×10^−6^*, *7.1×10^−7^*, and *3.4×10^−6^*, respectively. To investigate whether these identified associations were independent of the established GWAS-identified associations, we conducted conditional analyses by adjusting for the nearest GWAS-identified breast cancer risk variants. Conditional analyses showed that 24 genes from Exp-TWAS, 23 AS in 18 genes from spTWAS, and 4 APA in 4 genes from APA-WAS remained statistically significant at Bonferroni-corrected significance levels (**Table S2-S4**). In other words, 189 genes uncovered in this study are the likely target genes for 76 risk loci identified in previous GWAS.

**Figure 1.**
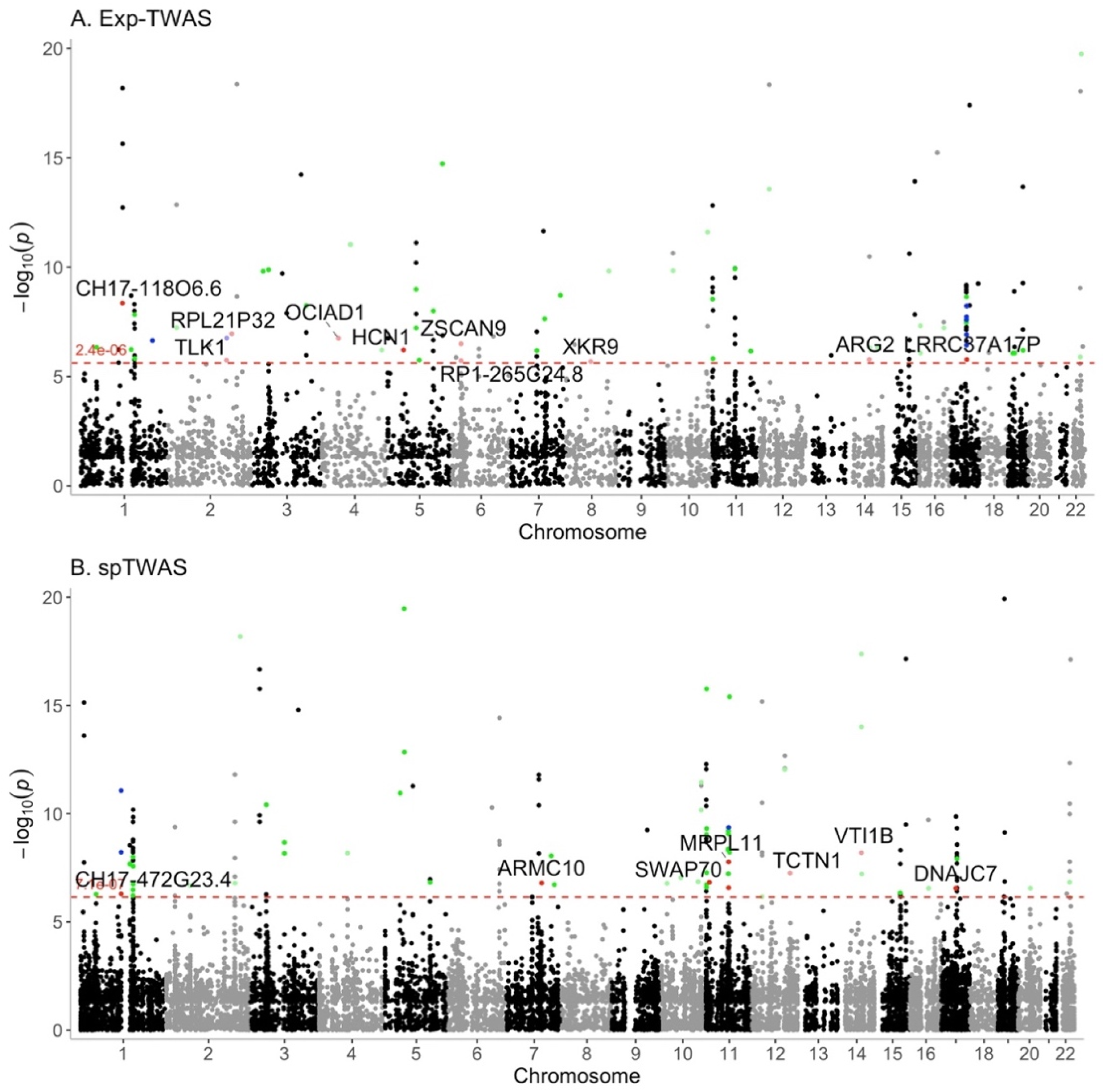
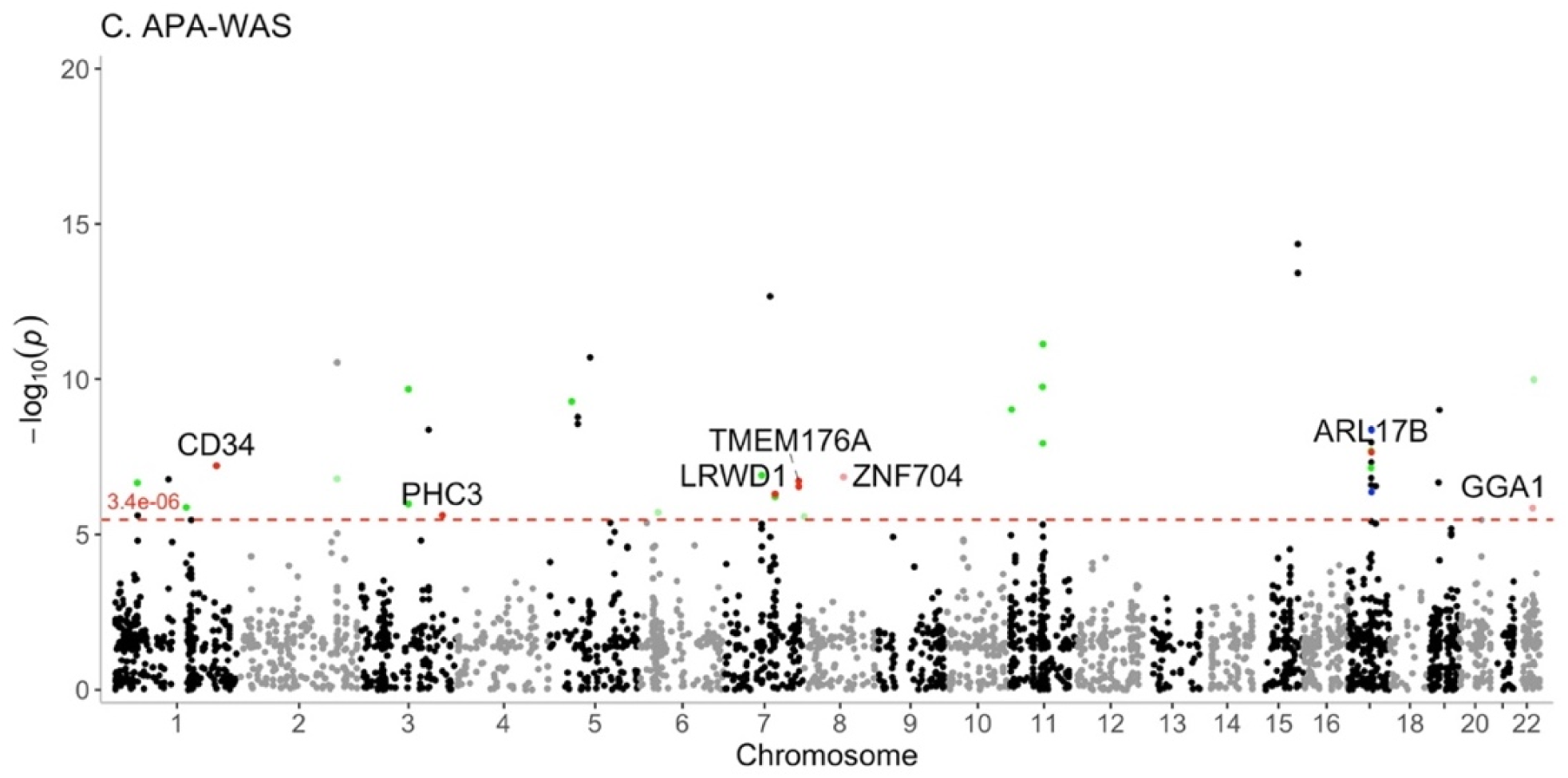
Manhattan plot for associations of genes identified with overall breast cancer risk. The red dotted lines represent the Bonferroni-corrected significance levels for Exp-TWAS (2.4×10^−6^), spTWAS (7.1×10^−7^), and APA-WAS (3.4×10^−6^). Red labeled dots and gene names highlight the significantly associated genes which have not been reported previously and are not located in any previous GWAS-identified loci, green dots indicate the significantly associated genes which have not been reported in any previous TWAS, and blue dots represent the significantly associated genes located at least 500Kb away from any previously GWAS-identified loci.

In total, we identified 263 genes (148 from Exp-TWAS, 106 from spTWAS, and 46 from APA-WAS, **Figure 2**) located at 87 unique loci associated with overall breast cancer risk at Bonferroni-corrected significance levels. Among these, 91 genes were not reported in any previous TWAS (**Table S2-S4**), and 20 are located at least 500Kb away from any previously GWAS-identified risk variants (**Table 1**).

**Figure 2.**
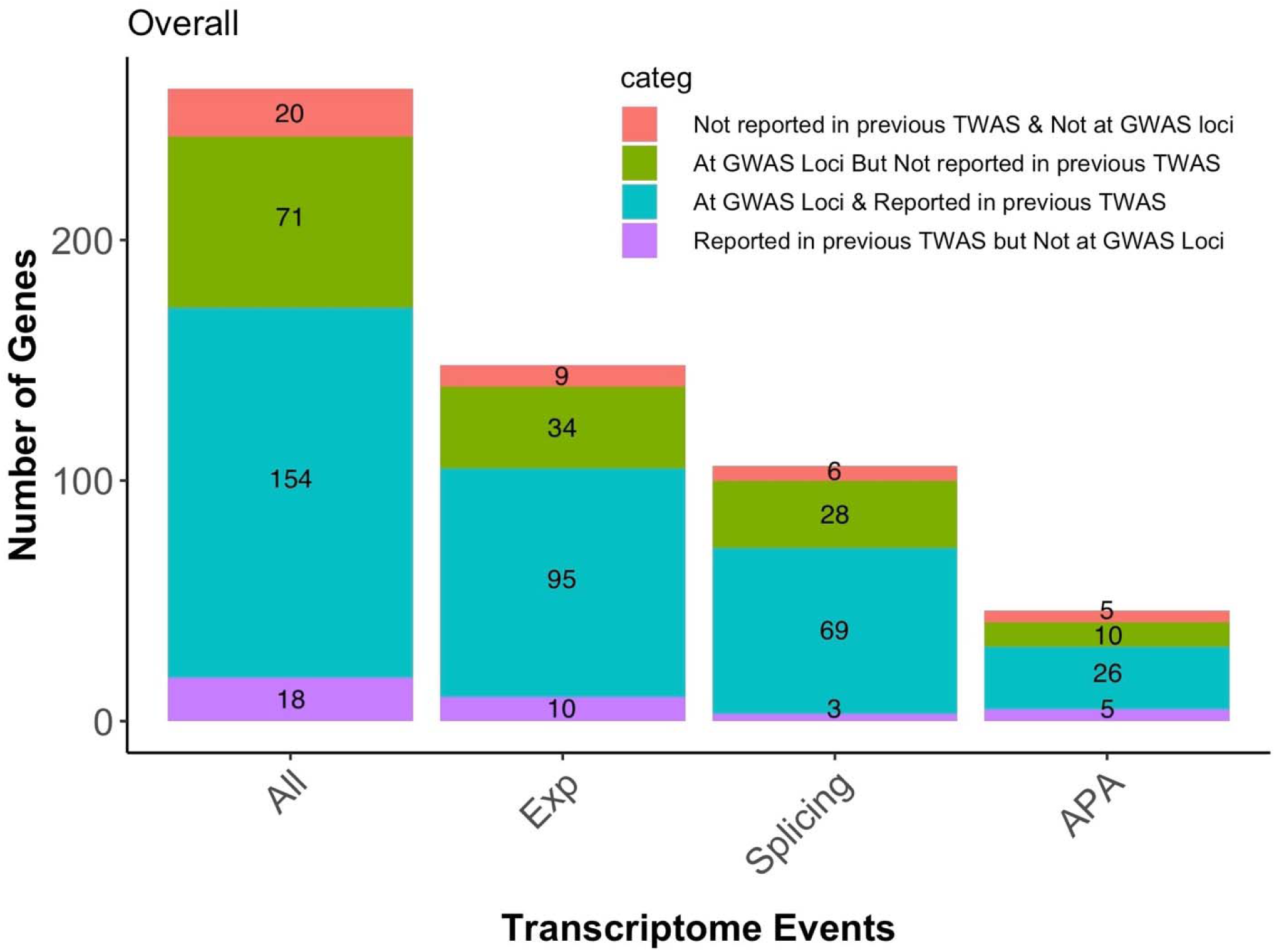
Bar plot of genes significantly associated with overall breast cancer risk identified in TWAS of different transcriptomic events.

**Table 1.** Newly identified breast cancer-associated genes not located in any GWAS risk loci and not reported in previous TWAS.

| Loci | Gene | Gene Type | Z Score | P Value | Identified by types of TWAS |
| --- | --- | --- | --- | --- | --- |
| 1p11.2 | CH17-472G23.4 | pseudogene | 5.02 | $5.06 \times 10^{-7}$ | AS |
| 2q31.1 | TLK1 | protein coding | 4.78 | $1.78 \times 10^{-6}$ | Expression |
| 2q32.1 | RPL21P32 | pseudogene | -5.20 | $1.96 \times 10^{-7}$ | Expression |
| 3q26.2 | PHC3 | protein coding | -4.72 | $2.42 \times 10^{-6}$ | APA |
| 4p11 | OCIAD1 | protein coding | -5.22 | $1.76 \times 10^{-7}$ | Expression |
| 5p12 | HCN1 | protein coding | 4.75 | $2.05 \times 10^{-6}$ | Expression |
| 6p22.1 | RP1-265C24.8 | lincRNA | 4.77 | $1.86 \times 10^{-6}$ | Expression |
| 7q22.1 | LRWD1 | protein coding | -5.03 | $5.00 \times 10^{-7}$ | APA |
| 7q22.1 | ARMC10 | protein coding | 5.25 | $1.55 \times 10^{-7}$ | AS |
| 7q36.1 | TMEM176A | protein coding | 5.21 | $1.88 \times 10^{-7}$ | APA |
| 8q13.3 | XKR9 | protein coding | -4.75 | $2.04 \times 10^{-6}$ | Expression |
| 8q21.13 | ZNF704 | protein coding | -5.27 | $1.40 \times 10^{-7}$ | APA |
| 11p15.4 | SWAP70 | protein coding | 5.26 | $1.44 \times 10^{-7}$ | AS |
| 11q13.2 | MRPL11 | protein coding | -5.63 | $1.84 \times 10^{-8}$ | AS |
| 12q24.11 | TCTN1 | protein coding | 5.44 | $5.43 \times 10^{-8}$ | AS |
| 14q24.1 | ARG2 | protein coding | -4.79 | $1.64 \times 10^{-6}$ | Expression |
| 14q24.1 | VTI1B | protein coding | -5.81 | $6.29 \times 10^{-9}$ | AS |
| 17q21.2 | DNAJC7 | protein coding | 5.14 | $2.75 \times 10^{-7}$ | AS |
| 17q21.31 | ARL17B | protein coding | 5.36 | $8.32 \times 10^{-8}$ | APA |
| 17q21.32 | LRRC37A17P | pseudogene | 4.79 | $1.65 \times 10^{-6}$ | Expression |
| 22q13.1 | GGA1 | protein coding | -4.65 | $3.39 \times 10^{-6}$ | APA |

We conducted similar analyses by estrogen receptor (ER) status and uncovered 14 additional genes associated with ER-positive breast cancer and 13 genes associated with ER-negative breast cancer at Bonferroni-corrected significance levels (**Table S5-S7**). Of these, 15 genes were not reported in any previous TWAS, and 6 genes are located at least 500Kb away from any previous GWAS-identified risk variant (**Table 2**). Of the 192 significantly associated genes identified by ER status, 39 genes showed a significantly different association by ER status at a Bonferroni-corrected *P<2.6×10^−4^* (0.05/192, **Table 3**), in which 26 genes showed an exclusive association with ER-positive breast cancer risk, while 11 genes showed an exclusive association with ER-negative breast cancer risk. Further conditional analyses showed that 40 genes including two additional genes *RHOD* from Exp-TWAS and *ZNF585B* from spTWAS by ER status remained statistically significant at Bonferroni-corrected significance levels (**Table S5-S7**). In other words, we identified likely 152 target genes for 69 risk loci reported from GWAS.

**Table 2.** Genes identified in analyses by ER status not located in any GWAS risk loci and not reported in previous TWAS.

| Loci | Gene | Gene Type | ER Positive |  | ER Negative |  | P for Het | Identified by types of TWAS |
| --- | --- | --- | --- | --- | --- | --- | --- | --- |
|  |  |  | Z Score | P Value | Z Score | P Value |  |  |
| 4q22.3 | SMARCAD1 | protein coding | -5.26 | $1.48 \times 10^{-7}$ | -2.25 | 0.02 | 0.06 | AS |
| 11q13.2 | C11orf80 | protein coding | 4.92 | $8.76 \times 10^{-7}$ | 1.78 | 0.07 | 0.23 | APA |
| 11q13.2 | KDM2A | protein coding | -4.65 | $3.34 \times 10^{-6}$ | -0.44 | 0.66 | $1.96 \times 10^{-5}$ | APA |
| 19p13.11 | CTD-2013N17.1 | pseudogene | -1.04 | 0.30 | 6.41 | $1.42 \times 10^{-10}$ | $3.02 \times 10^{-9}$ | Expression |
| 19q13.12 | ZNF585B | protein coding | 5.00 | $5.76 \times 10^{-7}$ | 2.23 | 0.03 | 0.26 | AS |
| 21q21.1 | CXADR | protein coding | -5.11 | $3.18 \times 10^{-7}$ | -2.00 | 0.05 | 0.27 | Expression |

**Table 3.**
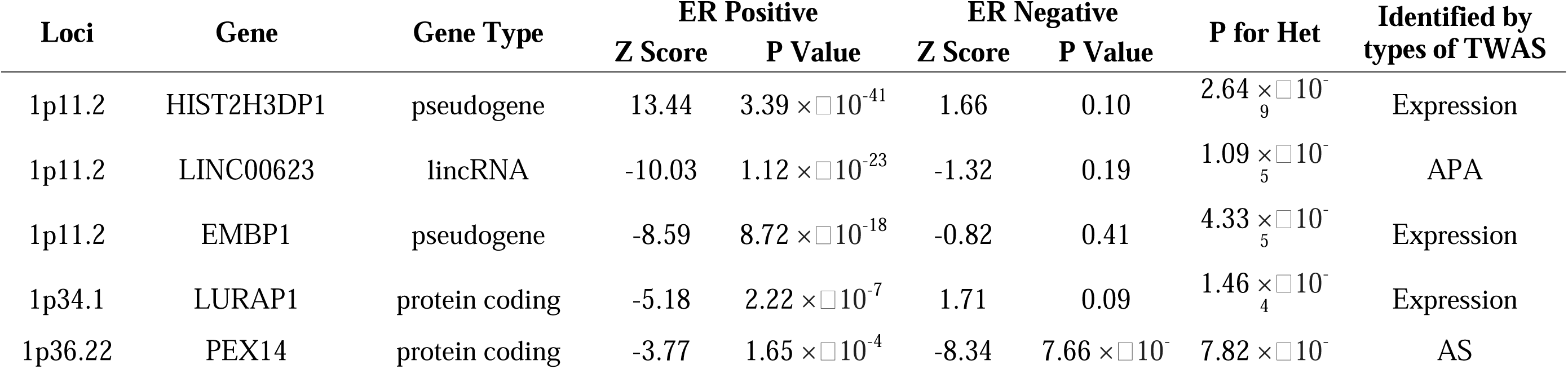
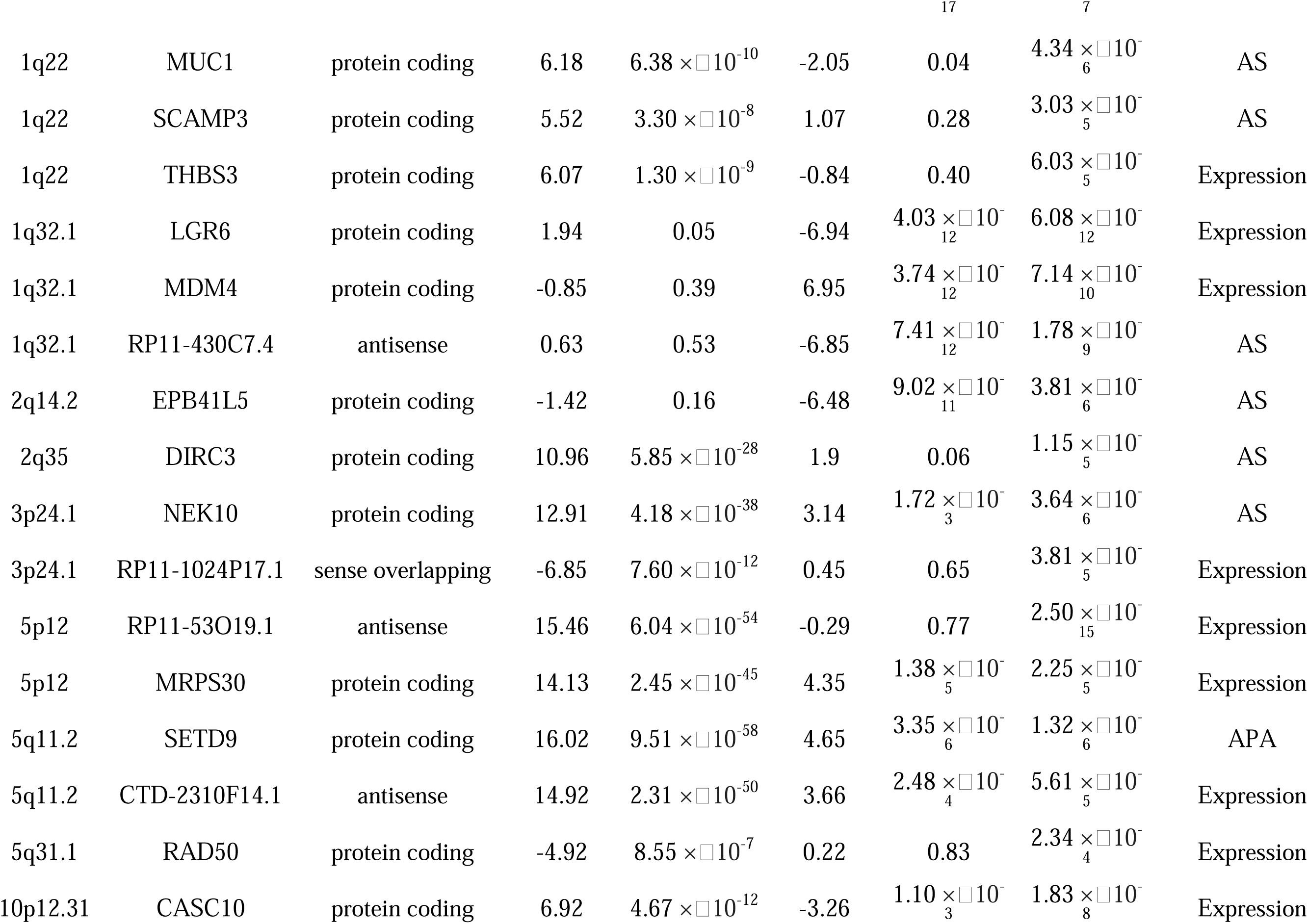
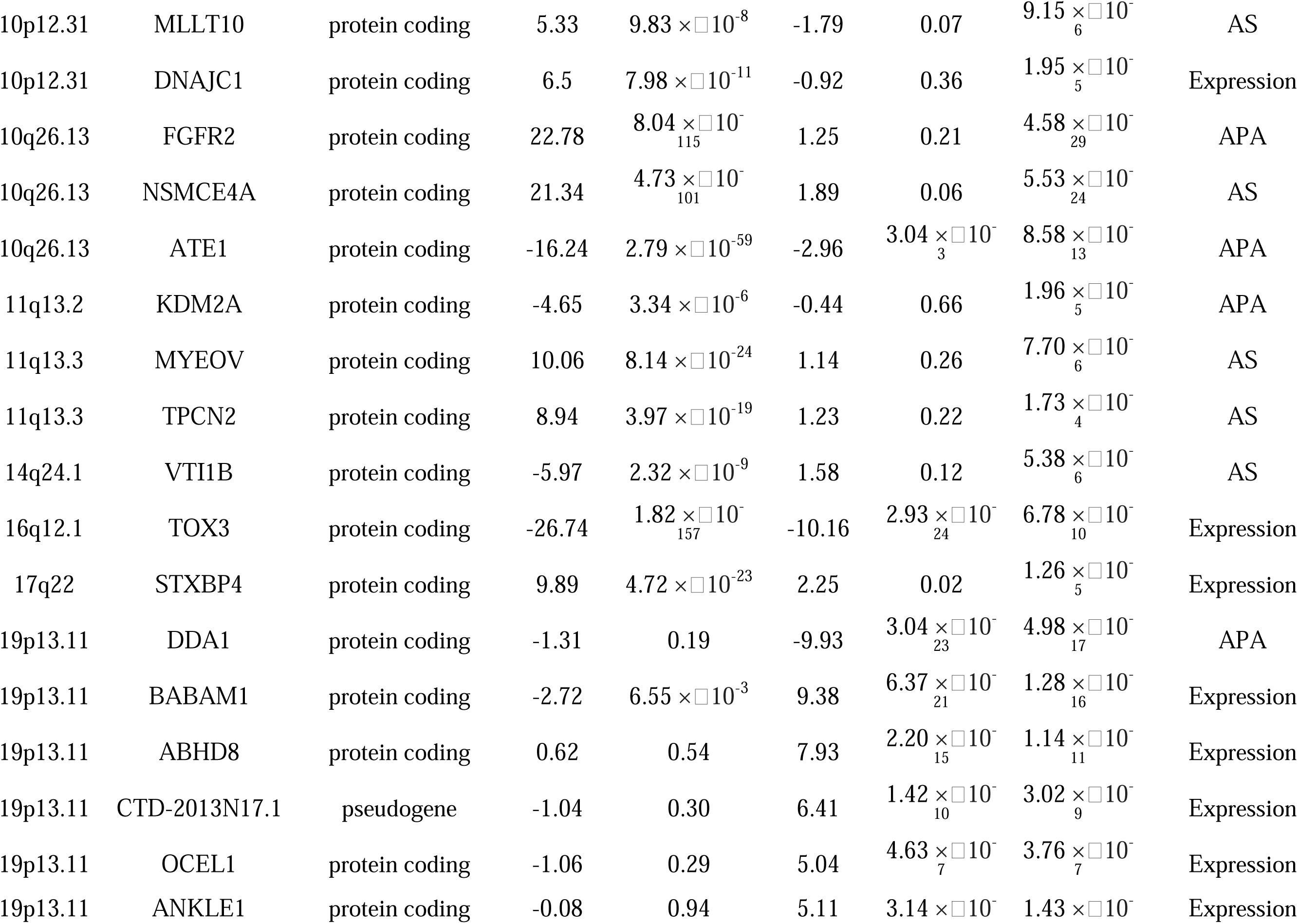
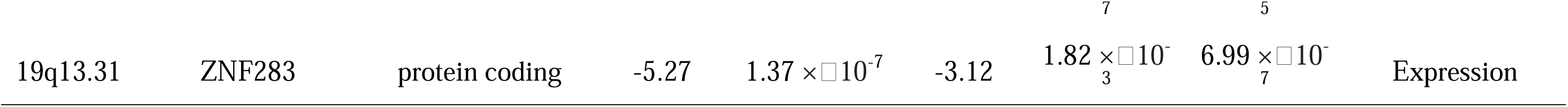
Genes showing significantly different associations by ER status of breast cancer.s.

In total, 290 genes were identified to be significantly associated with breast cancer risk, including 263, 14, and 13 genes associated with risk of overall breast cancer, ER-positive breast cancer, and ER-negative breast cancer, respectively. Of these, 103 genes were not reported in any previous TWAS and 46 genes are located at least 500kb away from any of the previously GWAS-identified risk loci. In conditional analyses, we showed that the associations for 196 genes were no longer statistically significant after adjusting for nearby GWAS-identified risk variants, suggesting that these genes are likely the targets for these risk variants, while for 48 genes for associations remained statistically significant at Bonferroni-corrected significance levels, suggesting that they may represent independent association signals.

To place our findings in the context of previous TWAS reports, we systematically evaluated results from earlier studies,^21–29^ including those conducted by our group.^2,15–20^ In total, significant associations for 653 genes have been reported in these previous studies in relation to breast cancer risk (**Table S13**), and these genes are mapped to 166 unique loci (**Table S11**). We found significant associations for genes located in 80 of these loci at the Bonferroni corrected significant level (**Table S11**). Of the remaining 86 loci, we identified at least one significantly associated gene in 59 loci at *P<0.001* (**Table S11**). Therefore, in addition to reporting novel findings, we replicated many earlier findings and provided further evidence supporting the likelihood that these genes are involved in breast cancer susceptibility.

We conducted KEGG pathway analyses including 216 protein-coding genes of the 290 genes identified in our study in relation to breast cancer risk. Presented in Table S8 are pathways enriched by these genes at a nominal *P<0.05* in analyses for all breast cancer combined or by ER status (**Table S8**). Several significantly enriched pathways in these analyses are well recognized to play a significant role in breast carcinogenesis primarily based on evidence from analyzing somatic alterations in breast cancer tissues, such as p53 signaling, homologous recombination, estrogen signaling, apoptosis, and PI3K-Akt signaling pathways. Several new pathways were also suggested in our analyses, including endocrine resistance, phospholipase D signaling and Rap1 signaling pathways.

We investigated the role of gene function in cell proliferation through *in vitro* experiments using breast cancer cell lines. Among the 216 protein-coding genes identified in our study, 203 genes had available CERES data derived from CRISPR-Cas9 which estimated the knockout effect of the genes across 44 invasive breast carcinoma cell lines. Of these, 22 genes had a median value of CERES score ≤*-0.5* (**Figure 3**), indicating their essential role in the proliferation of breast cancer cells. To further evaluate the functional significance of these genes in breast cancer risk, we conducted *in vitro* assays to determine whether gene knockdown using small interfering RNA (siRNA) may influence cell proliferation. Of the 216 protein-coding genes included in this assay, 93 genes showed a significant decrease in cell proliferation following knockdown compared to the siRNA scrambled negative control group (*P<0.05*; **Table S9**).

**Figure 3.**
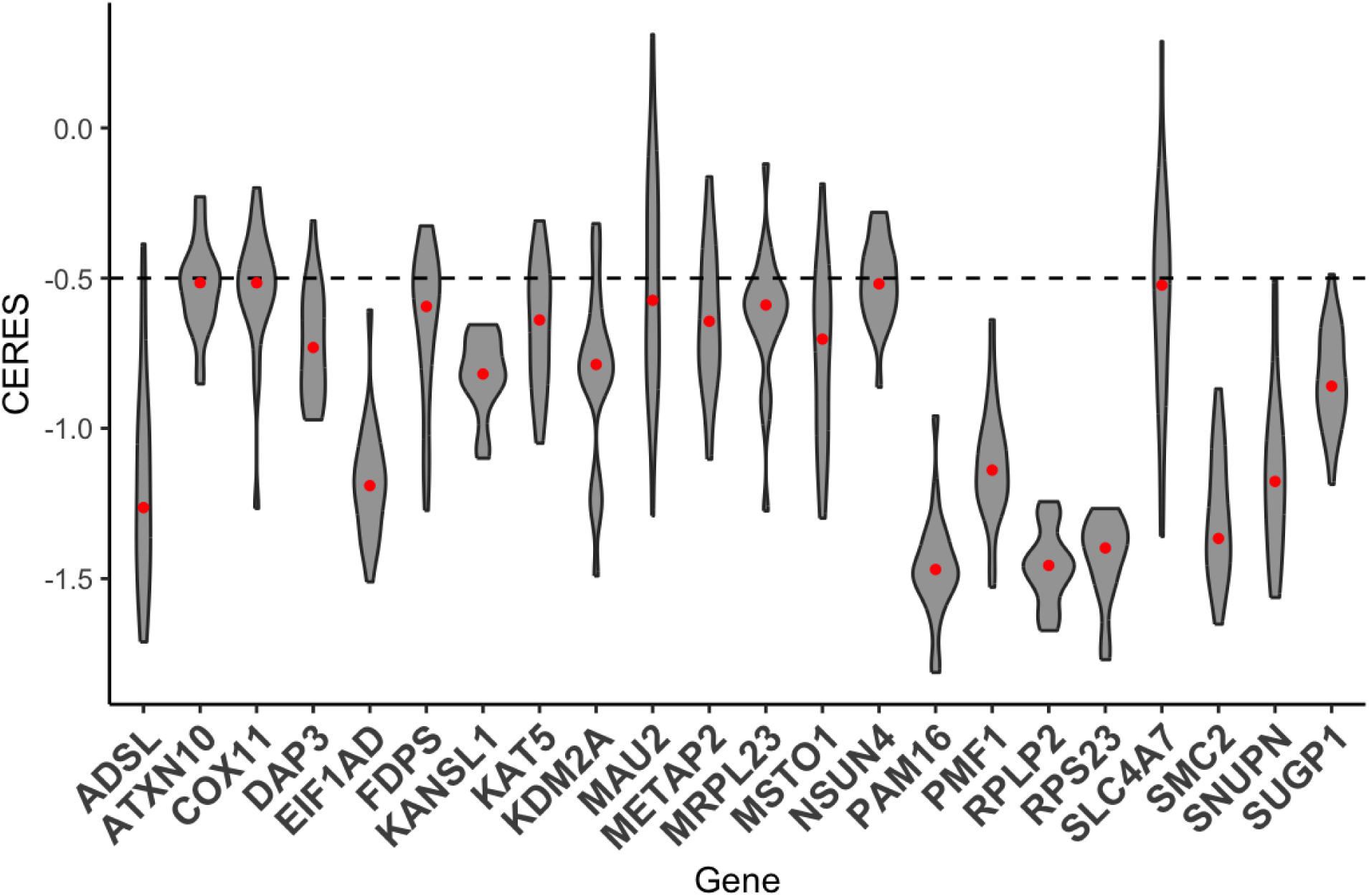
Essentiality of 22 selected putative breast cancer susceptibility genes from gene knockout experiments.

Furthermore, we investigated whether these putative breast cancer susceptibility genes identified in this study vary across different cell types in normal breast tissue. We performed differential gene expression analyses using single-cell RNA-Seq data from normal breast tissues of the GTEx project.^37^ Of the 203 protein-coding genes tested, 130 showed significantly differential expression levels in epithelial cell type compared to other cell types in normal breast tissue (Bonferroni-corrected *P<0.05*; **Table S10**), including 8 genes that are not reported in any previous TWAS and located at least 500kb away from any of the previously GWAS-identified risk loci.

## Discussion

Unlike many previous TWAS, we generated and used large amounts of transcriptome data in normal breast tissues for TWAS as these data are most relevant to breast cancer etiology. In this largest TWAS ever conducted to date, we systematically evaluated gene expression, AS, and APA to identify putative breast cancer susceptibility genes. In total, we identified 290 genes significantly associated with overall breast cancer risk (n=263) or cancer by ER subtypes (n=27). Of these, 103 were not reported in any previous TWAS and 46 are located at least 500Kb away from any previous GWAS-identified risk variants. Furthermore, we uncovered the likely target genes for 86 breast cancer risk loci identified in previous GWAS. These genes are involved in multiple cancer-related pathways, including several key pathways that are well recognized to play a significant role in breast carcinogenesis primarily based on evidence from analyzing somatic alterations in breast cancer tissues. These results suggest that genes driving breast tumor development might also influence an individual’s susceptibility to breast cancer. Our study provided substantial novel information on the potential mechanisms underlying breast cancer risk and significantly expanded the catalog of candidate breast cancer susceptibility genes.

This is the first breast cancer TWAS that included female participants from all three major ancestry populations (African, Asian, and European), which not only allowed us to evaluate the generalizability of TWAS findings across different populations but also enabled the identification of associations that are difficult to uncover in any single ancestry population. Also importantly, findings for a consistent association across ancestry groups support a causal association of the gene with breast cancer risk. For example, despite different genetic architectures across ancestry populations and thus different sets of genetic variants were used to predict transcriptomic events in each ancestry group, 46 genes were identified in association with breast cancer risk across all three populations and 70 genes were identified in two different populations, including several genes not located in any GWAS-identified risk loci. These data provided strong support for these genes as breast cancer susceptibility genes in these loci.

In addition to gene expression, we also evaluated two other transcriptomic events, splicing and APA, that were not adequately investigated in previous studies. Notably, 115 genes with Bonferroni-corrected significant associations were only identified through spTWAS or APA-WAS, including 17 new genes not reported before and not located in any previous GWAS-identified risk loci. Since spTWAS and APA-WAS focus on genetic variants involved in post-transcriptional regulation, these variants may influence breast cancer risk through regulatory mechanisms at the post-transcriptional level, which cannot be identified in traditional Exp-TWAS, which only considers gene expression. For example, the *RAD51B* gene has been implicated in breast cancer risk but not yet included in genetic testing for breast cancer.^38^ This gene was identified in our study using spTWAS, providing supports to formally classify this gene as a breast cancer susceptibility gene for future genetic testing.

Because of the large sample size for model building and the inclusion of models for three different transcriptomic events, we were able to identify 103 genes not yet reported in any previous TWAS studies, including some that are not even located in any previous GWAS-identified loci. Potential etiologic roles in breast cancer for some of these genes are supported by previous studies. For example, *MRPL11*, an essential protein of the large subunit of the mitochondrial ribosome, is frequently altered in tumors. High expression of *MRPL11* in stage 4 neuroblastoma was found to be correlated with poor prognosis. Abnormal expression of the *HCN1* gene was found previously to be significantly correlated with poor survival in breast cancer.^39^ *TLK1* (Tousled-like kinase 1) may have an important role in motility and metastasis and its inhibition chemosensitizes breast cancer (MDA-MB-231 and 4T1) cells to the drug doxorubicin.^40,41^ The *PHC3* gene was found to be a potential therapeutic target for epithelial tumors, including breast cancer.^42^ These previous studies provide some supports for potential roles of genes identified in our study in the etiology of breast cancer.

Although this is the largest TWAS ever conducted, the sample size for African and Asian-ancestry females remains small in association analyses. Therefore, some of the associations identified in the analysis of European-ancestry samples were not replicated in African and Asian-ancestry populations. Increasing the sample size in these two under-studied populations is warranted in future studies.

In summary, this large multi-ancestry TWAS identified 290 genes associated with breast cancer risk. Using data from diverse ancestries made it possible to uncover associations that would be difficult to detect in any single ancestry population. Additionally, incorporating alternative splicing and APA in our study allowed us to investigate post-transcriptional regulation, which significantly expanded the scope of the study. The findings of this study provided a comprehensive view into the genetics and biology of breast cancer.

## Data Availability

The genotyping and RNA-Seq data of samples from the Komen Tissue Bank generated in this study are available in the dbGaP database under accession code phs003535 [https://www.ncbi.nlm.nih.gov/projects/gap/cgi-bin/study.cgi?study_id=phs003535]. Summary-level statistics data from AABC are available in the GWAS Catalog (GCST90429846 for overall breast cancer, GCST90429847 for ER+ breast cancer, and GCST90429848 for ER− breast cancer). Summary-level statistics data from AABCG are available in the GWAS Catalog (GCST90296719 for overall breast cancer, GCST90296720 for ER+ breast cancer, and GCST90296721 for ER− breast cancer). Summary-level statistics data from BCAC are available in the GWAS Catalog (GCST010098 for overall breast cancer, GCST004988 for ER+ breast cancer, and GCST005076 for ER− breast cancer). GENCODE datasets are available from https://www.gencodegenes.org/human/release_26.html. All GTEx data are publicly available through dbGaP: phs000424.v8.p2.

## Code Availability

The developed pipeline and main source R codes used in this work are available from the GitHub website: https://github.com/pingjie/BCMATWAS.

## Acknowledgments

The content is solely the responsibility of the authors and does not necessarily represent the official views of the funding agents. The funders had no role in study design, data collection and analysis, decision to publish, or preparation of the manuscript. This research was supported in part by the US National Institutes of Health grants R01CA235553, R01CA202981, R01CA148667, and R01CA124558. Sample preparation and genotyping assays at Vanderbilt were conducted at the Survey and Biospecimen Shared Resources and Vanderbilt Technologies for Advanced Genomics, which are supported in part by the Vanderbilt-Ingram Cancer Center (P30CA068485). Data analyses were conducted using the Advanced Computing Center for Research and Education (ACCRE) at Vanderbilt University. Biospecimens from the Susan G. Komen Tissue Bank at the IU Simon Cancer Center were used in this study. We thank contributors, including those from Indiana University, who collected data used in this study, as well as donors and their families, whose help and participation made this work possible. Additional information, including grant support information for participating studies of the ABCC and AABCG, is provided in the Supplementary Note.

## Author Contributions

J.P. and W.Z. conceived and designed the study. Q.C., S.A., M.E.B., Y.C., J.-Y.C., Y.-T.G., M.G.-C., J.G., J.J.H., M.I., E.M.J., S.-S.K., C.I.L., K.Matsuda, K.Matsuo, K.L.N., B.N., O.I.O., T.P., S.K.P., B.P., M.F.P., M.S., D.P.S., M.A.T., S.Y., Y.Z., T.A., A.M.B., A.F., A.J.M.H, H.I., M.K., E.-S.L., T.M., B.S.E.M., D.-Y.N., K.M.O., O.O., A.F.O., M.-H.P., S.R., T.Y., E.N.B., M.H., S.-K.L., J.O., C.R.W, M.L.C., C.B.A., D.H., D.K., J.R.P., X.-O.S., C.A.H., J.L., and W.Z. recruited the study participants, and collected the data and specimens. J.P., G.J., Q.C., J.W., J.-Y.C., Y.-T.G., M.G.-C., J.G., M.I., E.M.J., S.-S.W., K.Matsuda, K.Matsuo, S.K.P., B.P., M.A.T., S.Y., Y.Z., T.A., H.I., M.K., E.-S.L., D.-Y.N., A.F.O., M.-H.P., T.Y., E.N.B., M.H., S.-K.L., C.R.W, H.Zhang, H.Zhao, M.L.C., C.B.A., D.H., D.K., X.-O.S., and X.G. managed sample and data preparation, or carried out quality control. J.P., G.J., Z.C., R.T., J.B., Y.X., B.L., and X.G. analyzed the data. J.P., G.J., Z.C., B.L., X.-O.S., and W.Z. interpreted the findings. J.P., Z.C., R.T., Y.X., H.Zhang, H.Zhao, J.R.P., C.A.H., X.G., J.L., and W.Z. drafted or substantively revised the paper.

## Competing Interests

Olufunmilayo I. Olopade (O.I.O.) is co-founder at CancerIQ, serves as Scientific Advisor at Tempus. Michael F. Press (M.F.P.) declares advisory board membership for Agilent; honoraria from Physicians’ Education Resource, LLC (PER) and Medscape; ownership interest in TORL Biotherapeutics; and fee-for-service agreements from 1200 Pharma, Ambrx, AstraZeneca, Inc., Novartis Pharmaceuticals Corporation, TORL Biotherapeutics, TRIO, TRIO-US, and Zymeworks. All other authors declare no competing interests.

## Methods

### Study populations

The overall genetic prediction models of gene expression, APA, and exon junction events were built using normal breast tissue data from three resources: the Susan G. Komen Normal Tissue Bank, the Asia Breast Cancer Consortium (ABCC), and the Genotype-Tissue Expression (GTEx) project.

Normal breast tissue samples donated by 182 European-ancestry, 49 Asian-ancestry, and 150 African-ancestry women to the Susan G. Komen Normal Tissue Bank, and 156 Asian-ancestry women from the ABCC were used in this study to generate new genomic and transcriptomic data to build genetic models to predict gene expression, APA, and splicing events, along with existing data from the GTEx project. In the Komen tissue bank, normal breast biopsies collected from the upper outer quadrant of the breasts were frozen within an average of six minutes from the time of biopsy, allowing for the preservation of the tissue quality.

Genomic DNA samples from all 381 females recruited in the Komen bank and 156 females in the ABCC were genotyped using the Illumina MEGA platform at Vanderbilt University Medical Center (VUMC). The other 73 Asian samples from the ABCC were genotyped using the Affymetrix Array platform. For model building, we excluded genetic variants with a minor allele frequency (MAF) < 5%, a Hardy-Weinberg equilibrium (HWE) p-value < 10^−4^, and a missing genotyping rate > 5%. SNPs with a consistency rate of < 98% among duplicate samples were also excluded. All genotyping data were imputed using the Trans-Omics for Precision Medicine (TOPMed) as a reference panel. Genetic variants with an imputation quality score of (r^2^) < 0.8 were excluded. Since it is difficult to determine the effect allele, multi-allelic SNPs and strand-ambiguous SNPs (with alleles A/T or C/G) were also excluded. Finally, SNPs not included in the final analysis dataset of the breast cancer GWAS data were excluded for model building.

Total RNA was extracted and purified using Qiagen’s AllPrep DNA/RNA/miRNA Universal Kit (Qiagen) following the manufacturer’s instructions. The quantity and quality of the DNA/RNA samples were checked by Nanodrop (E260/E280 and E260/E230 ratio) and by separation on an Agilent BioAnalyzer. Rnase H was used to remove rRNA. Each sample was sequenced pair-ended with a read length of 100 bp using DNBSEQ on BGISeq. A minimum of 10M reads were obtained for each sample.

RNA sequencing and whole-genome sequencing data of the GTEx project for normal breast tissues from European-ancestry women were downloaded from the dbGap (Accession Number: phs00424.v8.p2).

### GWAS summary statistics data for breast cancer risk

In this study, we conducted cross-ancestry multi-study TWAS using GWAS summary statistics data from three large breast cancer genetic research consortia: ABCC, AABCG, and BCAC. All studies were approved by relevant institutional ethical committees. The detailed descriptions of participating studies are described in the Supplementary Information.

In brief, the 133,384 individuals with breast cancer and 113,789 controls of European ancestry included in this analysis were from the BCAC, which consisted of three datasets: iCOGS (38,349 individuals with breast cancer and 37,818 controls), OncoArray (80,125 individuals with breast cancer and 58,383 controls), and other GWAS (14,910 individuals with breast cancer and 17,588 controls) (Table S1). For European-ancestry participants, we used summary statistics data generated in the BCAC. Individuals of Asian ancestry included in this analysis were 27,116 individuals with breast cancer and 112,407 controls recruited by studies in the AABC and the BCAC. For African-ancestry participants, we used summary statistics data generated in the AABCG that included 18,034 female cases and 22,104 female controls of African ancestry. A fixed-effects model was used for ancestry-specific meta-analyses and cross-ancestry meta-analyses for risk of overall breast cancer and estrogen receptor (ER) subtypes using METAL.

### Transcriptome data profiling and processing

#### Gene expression

RNA sequencing (RNA-Seq) data were processed following the mRNA analysis pipeline of the Genotype-Tissue Expression (GTEx) project.^37^ The raw data alignment to the human reference genome (hg38) was performed using a two-pass method of the Spliced Transcripts Alignment to a Reference (STAR) software.^43^ Coding gene and noncoding RNAs annotation in the human genome were based on GENCODE version 26.^44^ Gene-level expressions were quantified from the aligned BAM files using RNAseQC v1.1.9.^45^ Gene-level read counts and transcript per million (TPM) values were generated by applying the following read-level filters: 1) reads were uniquely mapped (with a mapping quality of 255 for STAR BAMs); 2) reads were aligned in proper pairs; 3) the read alignment distance was ≤ 6; 4) reads were fully contained within exon boundaries. Reads that overlapped with introns were not included in the counts. To normalize gene expression values for all samples from a single ancestral group in a study, the following procedure was applied: 1) read counts were normalized between samples using TMM; 2) genes were selected based on expression thresholds, requiring a minimum of ≥ 0.1 TPM in ≥ 20% of samples and ≥ 6 reads (unnormalized) in ≥ 20% of samples; 3) expression values for each gene were inverse normal transformed across samples.

#### APA events

The levels of APA events were quantified by estimating the Percentage of Distal polyA site Usage Index (PDUI) using DaPars v2.0 for each sample from the aligned RNA-Seq data.^46^ Any APA event with more than 5% missing values across all samples from a single ancestral group in a study was removed. To ensure comparability, quantile normalization was carried out to transform the quantified PDUI values of APA for each sample to the same distribution.

#### Splicing events

We quantified splicing based on the intron excision phenotypes computed by LeafCutter^47^ following the splicing quantification pipeline of the GTEx project. Briefly, intron usage and intron clusters were quantified and generated using LeafCutter. Introns with few counts or low complexity (diversity of counts across samples) were filtered out as follows to avoid numerical issues with the calculation of Beta-approximated empirical p-values in FastQTL: introns without any read counts in >50% of samples, or with fewer than max (10, 0.1n) unique values, where n is the sample size, were filtered out. Additionally, introns with insufficient variability across samples were removed based on thresholds applied to a z-score of clusters read fractions across individuals. The filtered counts were normalized using LeafCutter.

### PEER factors of gene expression variation

Probabilistic estimation of expression residuals (PEER) factors were calculated to correct for batch effects and other potential experimental confounders in further model building.^48,49^ The number of PEER factors was determined as a function of sample size as suggested in the GTEx protocol previously.^37^ The PEER factors were calculated for all samples from each ancestral group of each study.

### Genetic prediction model building

In this study, we built several genetic prediction models for each type of transcriptomic events (gene expression, APA events, and splicing events).

For expression data of the GTEx project, we used the joint-tissue imputation (JTI) approach, which borrows information across transcriptomes of different tissues to improve prediction performance. Besides breast tissue, data from 31 other tissues with both RNA-Seq and WGS data available with a sample size larger than 50 were borrowed in the JTI approach to leverage shared genetic regulation and improve prediction performance in a tissue-dependent manner. Gene expression levels were predicted using genetic variants within a flanking distance of ±500kb from the respective gene boundaries. Five-fold cross-validation was used to validate the models internally. Genes with a model prediction of *R^2^>0.01* (≥10% correlation between predicted and observed gene expression) were included for association analyses.

For expression data from Komen and the ABCC, as well as APA and splicing events from all three resources, all flanking genetic variants within a flanking distance of ±500Kb from the respective transcriptomic event were used to build the elastic net model. The model was implemented in the *glmnet* R package, with α *= 0.5*, as recommended by Gamazon *et al*.^50^ Age, PEER factors, and the top three genetic principal components were adjusted in model building. Five-fold cross-validation was used to validate the models internally. Prediction R values (the correlation between predicted and observed quantified level) were used to estimate the prediction performance of each prediction model.

In addition to building models using data from a single study, we also built models using data from multiple studies by using a modified version of the UTMOST (Unified Test for MOlecular SignaTures) framework.^35,36^ UTMOST is a framework for single and cross-tissue expression imputation. In this work, instead of cross-tissue, we applied the UTMOST method to the data of the same ancestral group from different resources, e.g., European data from GTEx and Komen.

The weights for SNPs in the prediction model were estimated with a LASSO penalty for both within- and cross-studies. Five-fold cross-validation tuned λ*_1_* and λ*_2_* for within- and cross-studies penalization. The updated UTMOST process ensured consistent hyperparameter pairs, selecting the pair with the lowest average error. Model performance was measured by predicted vs. observed expression correlation.

### Association analyses with breast cancer risk using GWAS data

Based on weight matrix and summary statistics data of SNPs from GWAS data, we evaluated the association between genetically predicted transcriptome events (gene expression, APA levels, or splicing levels) and breast cancer risk using the method from the S-PrediXcan tool.^51^ The details of the formula used in this method are

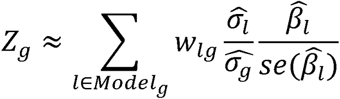

In brief, the Z-score was used to estimate the association between predicted transcriptome levels and breast cancer risk. In this formula, *w_lg_* is the weight of SNP *l* for predicting transcriptome levels of gene *g*. *β̂_l_* and *se* (*β̂_l_*) are the association regression coefficient and its standard error for SNP *l* in GWAS, and *σ̂_l_* and *σ̂_g_* are the estimated variances of SNP *l* and the predicted transcriptome levels of gene *g*, respectively. For this study, we estimated the correlations between SNPs included in the prediction models.

### Conditional Analyses

We additionally conducted conditional analyses by adjusting for the nearest GWAS-identified risk variants (the lead SNP, with the strongest association with breast cancer risk in the locus). Briefly, for each variant included in the model of genetically predicted transcriptomic events, GCTA-COJO analyses^52^ was performed to calculate the statistical significance with breast cancer risk after adjusting for the nearest lead variant. The resulting data were used for S-PrediXcan analyses to re-evaluate the associations conditioning on the nearest GWAS-identified risk variants.

### Functional enrichment analyses

A total of 216 putative protein-coding genes for breast cancer identified in this study were included for functional enrichment analyses. The WEB-based Gene Set Analysis Toolkit (WebGestalt) was used to perform for KEGG pathways enrichment analyses.^53,54^

### Knockout experiment data analyses

CRISPR-Cas9 knockout data in cell lines for invasive breast carcinoma were downloaded for the protein-coding genes identified in this study.^55^ The CERES score is a copy number-corrected measure of gene-targeting guide RNA depletion, with lower scores indicating a more essential role of the gene. Genes with a CERES score of ≤ −0.5 were considered as essential for the proliferation of breast cancer cells.^56^

We performed siRNA knockdown experiments in three breast cancer cell lines (MDA-MB-231, MCF-7 and T47D) for 216 genes identified in this study in association with breast cancer risk. Detailed methods are included in the Supplementary Note. For each gene, the viability score of the cells after siRNA knockdown was compared with the nontargeted siRNA controls (average viability of five nontargeted replicates per plate) using paired t-tests (two-sided, *P < 0.05* as cutoff). Genes associated with a decreased viability score after knockdown were considered to promote cell proliferation.

### Differential expression analysis of single-cell RNA-Seq data

We first converted the single-cell gene expression matrices from GTEx into count matrices. We normalized the unique molecular identifiers (UMIs) per cell to transcripts per 10,000 and then applied a log transformation to prepare the data for differential gene expression analysis. The top 2,000 most variable genes across all breast tissue samples were selected for the principal component (PC) analysis to determine the top 40 PCs. After removing batch effects, we used these top 40 PCs to construct a k-nearest neighbor graph of the cell profiles for eight different breast cell types, including adipocytes, lymphatic endothelial cells, vascular endothelial cells, luminal epithelial cells, fibroblasts, macrophages, myoepithelial cells, and pericytes. Finally, we used Wilcoxon’s rank-sum test (with a P-value cutoff of 0.05) to find differentially expressed genes between luminal epithelial cell type and all the others. Seurat v5.2.0 was used for all the single-cell RNA-Seq data analyses.^57^

## Notes

### Author Declarations

Ethics committee/IRB of Vanderbilt University Medical Center gave ethical approval for this work

