## Supplementary Figures for "Multi-Ancestry Transcriptome-wide Association Studies Uncover New Insights into Breast Cancer Genetics and Biology"

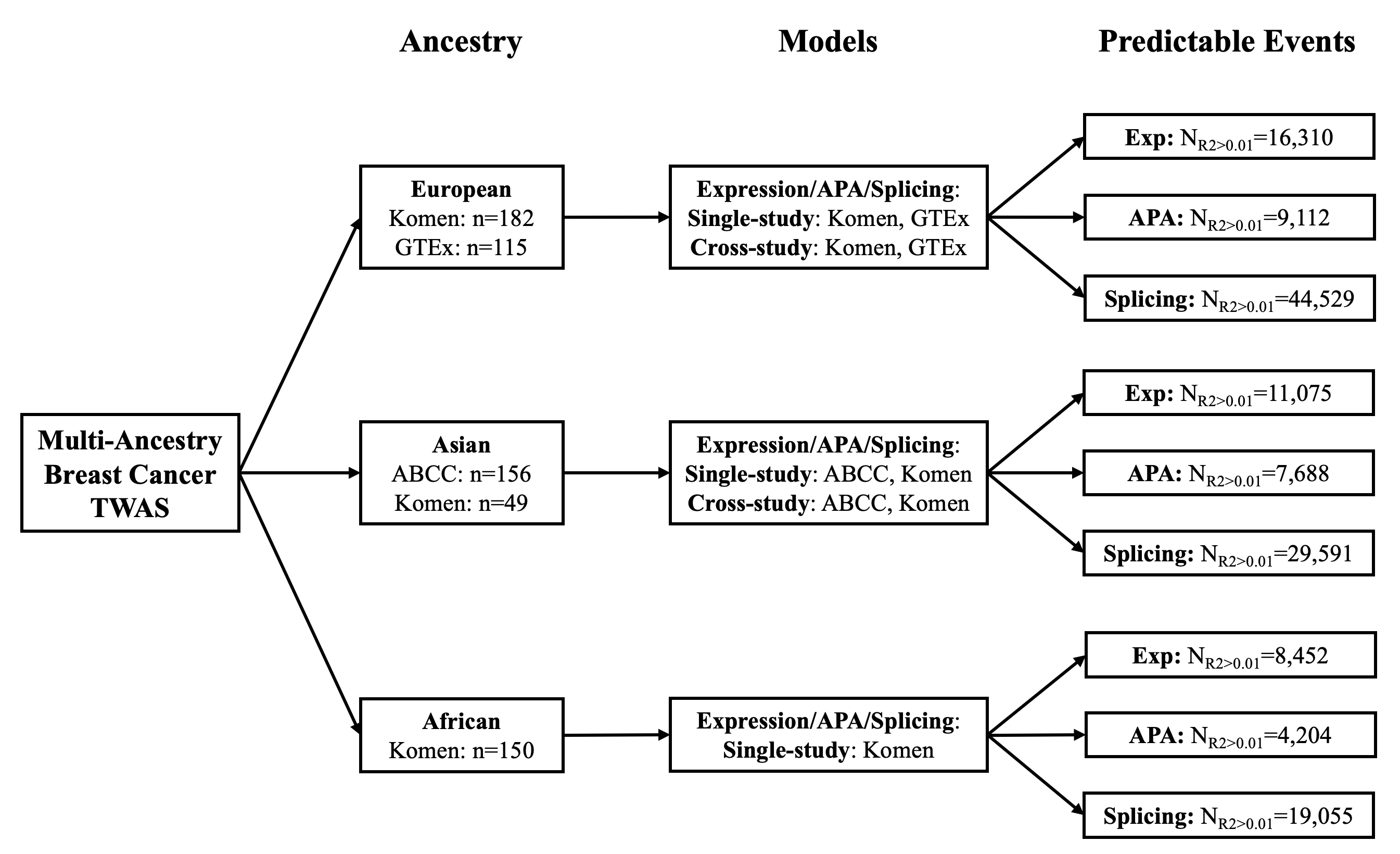

**Figure S1**. Summary of genetic model building for TWAS.

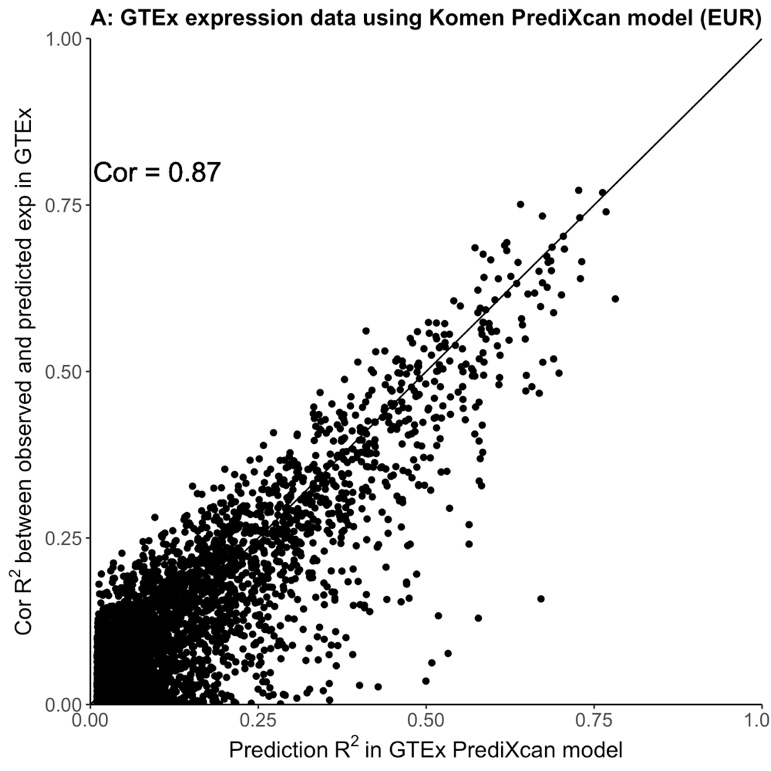

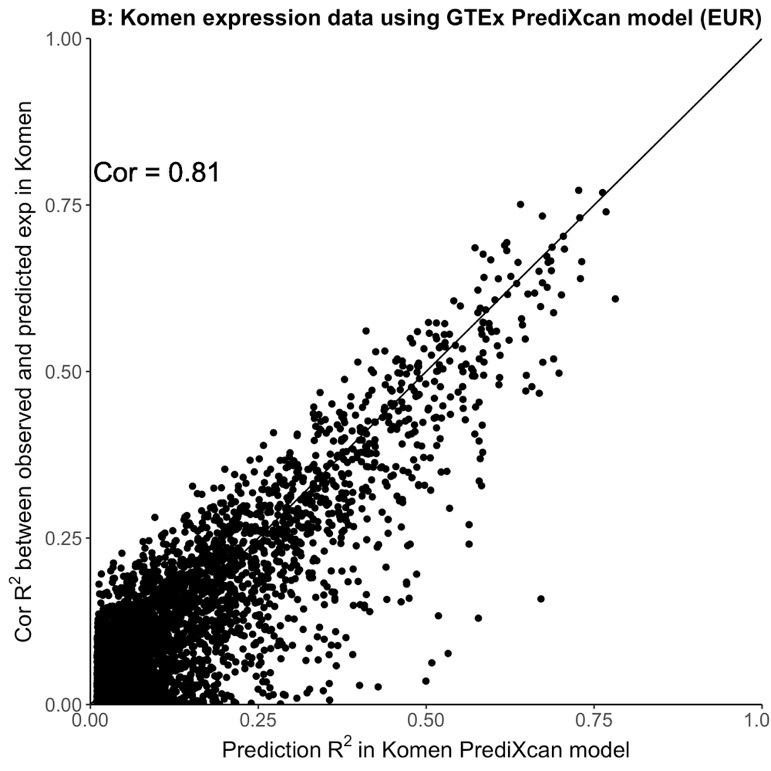

**Figure S2.** Model Validation. The x axis represents the prediction performance (R^2^) in GTEx dataset (A) and Komen dataset (B). The y axis represents the correlation coefficient R^2^ between observed expression and predicted expression using models from the other dataset (A: GTEx using Komen models; B: Komen using GTEx models). Each dot represents one gene. We predicted the expression level of each gene in one dataset (A: GTEx; B: Komen) using the model derived from the other dataset (A: Komen; B: GTEx) for that gene and compared the observed expression level of that gene in the first dataset to estimate a R^2^. We performed this correlation analysis for all genes that can be predicted with a R^2^ of 0.01 or higher in both datasets and showed a correlation coefficient of 0.87 and 0.81, supporting the validity of the models built for the study.
