## Supplementary Notes for "Multi-Ancestry Transcriptome-wide Association Studies Uncover New Insights into Breast Cancer Genetics and Biology"

### **Supplementary Methods**

#### **I.** **Description of participating studies in African American Breast Cancer Genetic (AABCG)**

The African American Breast Cancer Genetic (AABCG) consortium consists of whole genome sequencing data, newly generated genotyping data using the Multi-Ethnic Genotyping Array (MEGA), and genotyping data from existing studies/consortia. Relatives or the same participants (a close relationship with a Pi-HAT estimate >0.45) may have been genotyped by different arrays. For these, the samples genotyped by an array of a higher density were kept.

**1. Whole genome sequencing data**

The whole genome sequencing data included 1,337 breast cancer cases and 658 cancer-free controls (from five studies: SCCS, NBHS, STSBHS, GBHS, and MEC) which were sequenced using the Illunima HiSeq X Ten and BGIDEQ-500 platforms, and 71 cases and 1,639 controls from SCCS which were sequenced at >20x coverage on the Illumina HiSeq X and NovaSeq platforms with paired-end 150 bp reads. Details on sequencing library construction and data processing have been previously published^1^. In brief, all of the sequencing samples reached the average sequencing depth with at least 30X. The sequencing reads were aligned to the human reference genome (GRCh38) using the Burrows-Wheeler Aligner (BWA) program (version 0.79a). The mapped reads were further processed by removing the duplicated reads using MarkDuplicates from the Picard tool and recalibrating the base quality scores using BaseRecalibrator.

**1.1 Southern Community Cohort Study (SCCS)**

The SCCS is a prospective cohort study focused on the recruitment of a low-income, predominantly African-American population from a 12-state area of southeastern U.S^2,3^. Approximately 86,000 study participants aged 40–79 years were recruited between 2002 and 2009. About 32,500 of the SCCS participants were African American women. An in-person interview was completed at enrollment. Participants were asked to donate a 20-ml blood sample, and a buccal cell or saliva specimen was accepted if the subject did not wish to donate blood. To obtain follow-up data on cancer development, procedures for data linkage, processing, and quality control were established with the 12-state cancer registries covering the SCCS catchment area (Alabama, Arkansas, Florida, Georgia, Kentucky, Louisiana, Mississippi, North Carolina, South Carolina, Tennessee, Virginia, and West Virginia). In SCCS, samples from 321 cases and 376 controls were sequenced by Illunima HiSeq X Ten and BGIDEQ-500 platforms. In addition, 71 cases and 1,639 controls from SCCS were sequenced at >20x coverage on the Illumina HiSeq X and NovaSeq platforms with paired-end 150 bp reads, which were called as WGS-2 in this study.

**1.2 Nashville Breast Health Study (NBHS)**

The NBHS is a population-based case-control study conducted in the Nashville metropolitan area^4^. Participants were recruited between 2001 and 2011. Eligible cases were women newly diagnosed with primary breast cancer between 25 and 75 years of age and with no prior history of cancer other than nonmelanoma skin cancer. Breast cancer cases (n =2,694) were identified through the Tennessee State Cancer Registry and five major hospitals in Nashville that provide medical care for breast cancer patients. Controls (n =2,384) were identified via random digit dialing of households in the same geographic area as the cases. Saliva samples were collected as a source of genomic DNA for genetic studies of breast cancer. In NBHS, samples from 91 cases and 16 controls were sequenced by BGIDEQ-500 platform.

**1.3 Southern Tri-State Breast Health Study (STSBHS)**

The STSBHS is a population-based case-only study conducted in Tennessee, South Carolina, and Georgia^5^. It recruited women with incident invasive breast cancer diagnosed between ages 25 and 75 years from 2012 to 2018, without any prior cancer history other than non-melanoma skin cancer. Breast cancer cases were identified through the Tennessee Cancer Registry, the South Carolina Central Cancer Registry, and the Georgia Comprehensive Cancer Registry. In STSBHS, samples from 421 cases were sequenced by BGIDEQ-500 platform

**1.4 Multiethnic Cohort Study (MEC)**

The MEC is a prospective cohort study conducted in Hawaii and the Los Angeles area and included 215,251 study participants recruited between 1993 and 1996^6^. African American study participants were recruited from the Los Angeles area. Data collection at baseline included a detailed, self-administered questionnaire that obtained information on basic demographic variables and several lifestyle and medical variables that have been associated with cancer risk. Incident cancer cases were identified through two state-wide Surveillance, Epidemiology, and End Results (SEER) registries: the Hawaii Tumor Registry and the California State Cancer Registry. Blood samples were collected from a substantial portion of the cohort. In MEC, samples from 211 cases and 119 controls were sequenced by Illunima HiSeq X Ten.

**1.5 Ghana Breast Health Study (GBHS)**

The GBHS is a population-based case-control study conducted in Accra and Kumasi, Ghana, between 2013 and 2015^7,8^. Breast cancer cases were identified from women recommended for biopsy of a breast lesion suspicious for malignancy at one of the three major cancer treatment hospitals in Ghana (Korle Bu Teaching Hospital in Accra, and Komfo Anokye Teaching Hospital, and Peace and Love Hospital in Kumasi); or women presenting for treatment of pathologically-confirmed breast cancer at Korle Bu, Komfo Anokye, or Peace and Love Hospitals within one year of diagnosis. Controls were frequency matched to cases by age and district of residence. In GBHS, samples from 293 cases and 147 controls were sequenced by BGIDEQ-500 platform.

**2. Newly generated genotyping data using Multi-Ethnic Genotyping Array (MEGA)**

In the African American Breast Cancer Genetic (AABCG) consortium, we genotyped samples from multiple studies using the Multi-Ethnic Genotyping Array (MEGA). The MEGA chip contains over 2 million variants with an excellent genomic coverage of common variants across multi-racial populations. Samples were genotyped in Vanderbilt University Medical Center (SCCS, NBHS, STSBHS, MDABCS, CCPS, NBCS, NC-BCFR, NYUWHS), Roswell Park Comprehensive Cancer Center (BWHS, WCHS), and University of Southern California (MEC). Samples that overlapped between whole genome sequencing data and genotyping data were only kept in the whole genome sequencing dataset.

Quality control (QC) procedure includes: samples were excluded if they (i) were not genetically female; (ii) had a call rate <95%; (iii) had a low proportion of African ancestry (<25%) using Admixture^9^, using 1000 Genome samples as reference; or (iv) had a close relationship with a Pi-HAT estimate >0.45 (one pair was excluded). Variants were excluded if they had (i) a call rate <95%; (ii) a P value <10^-6^ in the Hardy-Weinberg equilibrium test among the controls; (iii) a consistent rate <98% across duplicated QC samples; or (iv) inconsistent alleles from 1000 Genome data.

**2.1 Southern Community Cohort Study (SCCS)**

The SCCS has been described above. After quality control, there were 708 cases and 678 controls from SCCS newly genotyped by MEGA. In addition, 2,265 controls from SCCS were selected for cases from NBHS, STSBHS, MDABCS, and NC-BCFR. The controls were frequently matched on age and state of residence (only for NBHS and STSBHS).

**2.2 Nashville Breast Health Study (NBHS)**

The NBHS has been described above. After quality control, new MEGA genotyping data included 138 cases from NBHS, and 147 controls matched from SCCS.

**2.3** **Southern Tri-State Breast Health Study (STSBHS)**

The STSBHS has been described above. After quality control, new MEGA genotyping data included 692 cases from STSBHS and 683 controls matched from SCCS.

**2.4 M.D. Anderson Breast Cancer Study (MDABCS)**

All breast cancer cases in MDABCS are newly registered, histologically confirmed breast cancer patients at MD Anderson Cancer Center^10^. Basic demographic and epidemiological information including smoking, alcohol, education, and family history data were collected as part of institutional patient history database. Clinical data were abstracted from electronic medical records by clinical coding specialists. DNA were extracted from residual blood samples and banked in the institutional Blood Specimen Research Resource. In the MEGA genotyping data, controls from SCCS were frequently matched on age. After quality control, new MEGA genotyping data included 1,294 cases from MDABCS and 1,222 controls matched from SCCS.

**2.5 Chicago Cancer Prone Study (CCPS)**

The CCPS is a hospital-based case-control study designed to investigate the genetics of young-onset breast cancer. Cases with histologically confirmed breast cancer were enrolled through the Cancer Risk Clinic at the University of Chicago. Young-onset cases and African Americans were oversampled. Controls were gender- and age-matched with cases and enrolled from patients who visited the same hospital and were willing to donate blood for genetic studies. After quality control, new MEGA genotyping data included 366 cases and 279 controls from CCPS.

**2.6 Nigerian Breast Cancer Study (NBCS)**

The NBCS is an ongoing case-control study of breast cancer in Ibadan, Nigeria initiated in 1998^11,12^. Breast cancer cases were 20 years or older, ascertained at the University College Hospital, Ibadan, which is the oldest tertiary hospital in Nigerian with a catchment population of approximate three million. Controls were recruited from a randomly selected community in one of the communities adjoining the hospital. The majority of the study subjects were Yoruba and Yoruba is one of the populations selected by the International HapMap Project to represent African continent. After quality control, new MEGA genotyping data included 695 cases and 376 controls from NBCS.

**2.7** **Northern California Breast Cancer Family Registry (NC-BCFR)**

Incident breast cancer cases included women aged <65 years, identified through the SEER cancer registries of the Greater San Francisco Bay Area (diagnoses 1995-2009) and the Sacramento region (diagnoses 2005-2006).^13,14^ All cases with indicators of inherited breast cancer were included. Among cases aged 35-64 years without such indicators, cases from racial and ethnic minority populations were oversampled. Population controls were identified through random-digit dialing and frequency matched to cases diagnosed from 1995-1998 on 5-year age group and race/ethnicity, at a ratio of one control per two cases. In the MEGA genotyping data, controls from SCCS were selected by frequency matching on age. After quality control, new MEGA genotyping data included 185 cases from NC-BCFR and 213 controls matched from SCCS.

**2.8 New York University Women’s Health Study (NYUWHS)**

The NYUWHS is a cohort study which enrolled 14,274 women aged 34 to 65 years attending Guttman Breast Diagnostic Institute in New York City for yearly screening from 1985 to 1991^15,16^. Self-administered questionnaires were used to collect demographic, medical, anthropometric, reproductive, and dietary. Non-fasting peripheral venous blood was drawn prior to breast examination and serum samples were stored at -80°C for subsequent biochemical analyses. Up until 1991, women who returned to the clinic for annual breast cancer screening were asked to donate blood at each of their visits. Cases were breast cancer patients arising from in the cohort, and controls were women selected from the same cohort who were not diagnosed with breast cancer and matched to cases on age and follow up time. After quality control, new MEGA genotyping data included 72 cases and 58 controls from NYUWHS.

**2.9 Women’s Circle of Health Study (WCHS)**

The WCHS is a case-control study established in 2003 in the New York City metropolitan areas, and beginning in 2006, from 10 counties in New Jersey^17^. Eligible cases included women who were diagnosed with invasive breast cancer between 20 and 75 years of age and self-identified as European-American or African-American. Controls were initially identified through random digit dialing and were matched to cases by self-reported race and 5-year age categories. From 2009-2012, controls were recruited through community events, particularly through churches^18^. After quality control, new MEGA genotyping data included 1,326 cases and 851 controls from WCHS.

**2.10 Black Women’s Health Study (BWHS)**

The BWHS is a prospective cohort study which recruited approximately 59,000 African American women, aged 21-60 years, from all regions of the United States in 1995^19^. Participants were enrolled by completing a postal health questionnaire and were followed by mail questionnaires every two years. DNA samples were obtained from BWHS participants (26,800 women) by the mouthwash-swish method with all samples stored in freezers at −80°C. After quality control, new MEGA genotyping data included 1,282 cases and 1,879 controls from BWHS.

**2.11 Multiethnic Cohort Study (MEC)**

The MEC has been described above. In the MEGA genotyping data, cases were from MEC and controls were from African American Eye Disease Study (AFEDS). The AFEDS is a population-based cohort study conducted from April 2014 to April 2018 including 6,347 African American adults, 40 years of age and older residing in 32 census tracts in and around Inglewood, CA within Los Angeles County. The participation rate for eligible residents who completed the clinical examination and home interview was 80% (6,347 of 7,957 eligible). While the study was conducted to fill the gaps in our understanding of vision health in African American adults, selection was independent of eye health. Biological samples included a blood draw and/or a saliva sample depending on the selection by the participant after informed consent was completed. Data on demographic and behavioral characteristics, medical and ocular history, insurance status and access to care were collected, and a comprehensive eye examination was conducted. Detailed methods have been published elsewhere.^20^ A random sample of female participants from the cohort were selected for inclusion in the AABCGS genetic study based on number of budgeted tests (N=969). Women with a self-reported history of breast cancer were excluded. The mean age of the genotyping set was 57.8 years (SD=10.0) with a minimum age of 40 and a maximum age of 83 years. Of the 969 participants, 4.3% had less than a high school education, 17.4% had a high school education, 38.5% had some college education, and 37% had a college degree or higher (2.8% not reported). After quality control, the MEGA genotyping data included 1,194 cases from MEC and 914 controls from AFEDS.

**3. African American Breast Cancer Epidemiology and Risk (AMBER) consortium**

The AMBER is a collaboration of four studies: the Women’s Circle of Health Study (WCHS), the Black Women’s Health Study (BWHS), the Carolina Breast Cancer Study (CBCS), and the Multiethnic Cohort Study (MEC) funded by the National Cancer Institute. In the AMBER phase 2, a total of 4,224 study samples were genotyped by MEGA chip and a set of custom variants selected from breast cancer candidate loci at the Center for Inherited Disease Research at Johns Hopkins University.

**3.1 Carolina Breast Cancer Study (CBCS)**

The CBCS is a population-based case-control study conducted in 24 counties of central and eastern North Carolina^21^. From 1993 to 2001, it recruited women aged between 20 and 74 years and diagnosed with invasive breast cancer. African-American women and women aged less than 50 years were oversampled. Cases were identified by rapid case ascertainment system in cooperation with the North Carolina Central Cancer Registry. Controls were selected from the North Carolina Division of Motor Vehicle (for women younger than 65 years) and United States Health Care Financing Administration (for women aged 65 and older). Controls were approximately frequency matched to cases by age and race. Blood samples were collected from participants with consent. After quality control, we included 602 cases and 1 control of African ancestry from CBCS in the AMBER phase2.

**3.2 Women’s Circle of Health Study (WCHS)**

The WCHS has been described above. After quality control, we included 472 cases and 243 controls of African ancestry from WCHS in the AMBER phase2.

**3.3** **Black Women’s Health Study (BWHS)**

The BWHS has been described above. After quality control, we included 307 cases and 2,098 controls of African ancestry from BWHS in the AMBER phase2.

**4. The GWAS of Breast Cancer in the African Diaspora (ROOT) consortium**

The ROOT consortium consists of samples from NBCS, BNCS, RVGBC, CCPS, BBCS, and SCCS. The samples in ROOT consortium were genotyped using the Illumina HumanOmni2.5-8v1 array.

**4.1 Baltimore Breast Cancer Study (BBCS)**

The BBCS is a case control study of breast cancer designed to identify and characterize markers of disease aggressiveness and poor outcome. From 1993 to 2003, incident breast cancer cases and controls were recruited from six hospitals in the greater Baltimore area, including the University of Maryland Medical Center, the Baltimore Veterans Affairs Medical Center, Union Memorial Hospital, Mercy Medical Center, and the Sinai Hospital. Controls were frequency matched to cases by race and age. After quality control, we included 94 cases and 102 controls of African ancestry from BBCS in the ROOT consortium.

**4.2 Barbados National Cancer Study (BNCS)**

The BNCS is a population-based case-control study of incident breast and prostate cancer in the predominantly African population of Barbados, West Indies^22^. Breast cancer cases were histologically confirmed incident cases identified through the only pathology department on the island, located at the Queen Elizabeth Hospital, between July 2002 and March 2006. Controls were selected from a national database provided by the Barbados Statistical Services Department, and were frequency matched to breast cancer cases at a 2:1 ratio and by 5-year age groups. Blood samples were collected from participants. After quality control, we included 92 cases and 227 controls of African ancestry from BNCS in the ROOT consortium.

**4.3 Racial Variability in Genotypic Determinants of Breast Cancer Risk Study (RVGBC)**

RVGBC is a hospital-based case-control study conducted in Philadelphia and Detroit metropolitan areas from 1999 to 2003. Breast cancer cases were identified in the University of Pennsylvania Health System and Karmanos Cancer Institute. Local advertisement was also put to recruit breast cancer cases living in the Philadelphia and Detroit area. Controls were recruited in the same way as cases except that they did not have breast cancer. Patients with breast cancer had to be diagnosed within 18 months of recruitment and have invasive ductal cancer. The study over-sampled women diagnosed with breast cancer under age of 40 years. After quality control, we included 143 cases and 254 controls of African ancestry from BNCS in the ROOT consortium.

**4.4 Chicago Cancer Prone Study (CCPS)**

The CCPS has been described above. After quality control, we included 365 cases and 376 controls of African ancestry from CCPS in the ROOT consortium.

**4.5 Nigerian Breast Cancer Study (NBCS)**

The NBCS has been described above. After quality control, we included 702 cases and 602 controls of African ancestry from NBCS in the ROOT consortium.

**4.6 Southern Community Cohort Study (SCCS)**

The SCCS has been described above. After quality control, we included 126 cases and 323 controls of African ancestry from SCCS in the ROOT consortium.

**5. The African American Breast Cancer (AABC) consortium**

The AABC consortium consists of samples from NBHS, NC-BCFR, CARE, CBCS, MEC, PLCO, SFBCS, WCHS, and WFBC. The samples in AABC consortium were genotyped using IlluminaHuman1M-Duo BeadChip.

**5.1 The Los Angeles component of the Women’s Contraceptive and Reproductive Experiences Study (CARE)**

The Women's CARE Study is a large multi-center population-based case-control study sponsored by the National Institute of Child Health and Human Development (NICHD)^23^. It was designed to examine the effects of oral contraceptive use on invasive breast cancer risk. Cases diagnosed with breast cancer between 34 and 64 years of age were recruited in five U.S. locations (Atlanta, Detroit, Los Angeles, Philadelphia, and Seattle). Cases in Los Angeles County were diagnosed from July 1, 1994 through April 30, 1998, and controls were sampled by random-digit dialing from the same population and time period. After quality control, we included 254 cases and 204 controls of African ancestry from CARE in the AABC consortium.

**5.2 The Prostate, Lung, Colorectal, and Ovarian Cancer Screening Trial (PLCO)**

The PLCO is a multicenter, two-armed, randomized trial designed to evaluate the screening efficacy for prostate, lung, colorectal and ovarian cancer^24^. It recruited approximately 155,000 men and women, aged 55-74 years, from 1993 to 2001. After quality control, we included 24 cases and 68 controls of African ancestry from PLCO in the AABC consortium.

**5.3 San Francisco Bay Area Breast Cancer Study (SFBCS)**

The SFBCS is a population-based case-control study of invasive breast cancer in Hispanic, African American and non-Hispanic White women in the San Francisco Bay Area^25^. Women aged 35–79 years, diagnosed with a first primary invasive breast cancer between 1995 and 2002 were identified through the California population-based Greater Bay Area Cancer Registry. Population controls were identified through random digit dialing, frequency matched to cases on race and ethnicity and 5-year age group. After quality control, we included 157 cases and 210 controls of African ancestry from SFBCS in the AABC consortium.

**5.4 Wake Forest University Breast Cancer Study (WFBC)**

The WFBC is a clinic-based case-control study at Wake Forest University Health Sciences from 1998 to 2008^26,27^. Incident breast cancer cases were recruited at the Wake Forest University Breast Care Center. Controls were recruited from the patient population receiving routine mammography at the Outpatient Radiology-Breast Screening Center. Blood samples (20 ml) were collected from all study subjects. After quality control, we included 113 cases and 138 controls of African ancestry from WGBC in the AABC consortium.

**5.5** **Nashville Breast Health Study (NBHS)**

The NBHS has been described above. After quality control, we included 255 cases and 161 controls of African ancestry from NBHS in the AABC consortium.

**5.6 Northern California Breast Cancer Family Registry (NC-BCFR)**

The NC-BCFR has been described above. After quality control, we included 383 cases and 48 controls of African ancestry from NC-BCFR in the AABC consortium.

**5.7 Carolina Breast Cancer Study (CBCS)**

The CBCS has been described above. After quality control, we included 614 cases and 570 controls of African ancestry from CBCS in the AABC consortium.

**5.8 Multiethnic Cohort Study (MEC)**

The MEC has been described above. After quality control, we included 578 cases and 888 controls of African ancestry from MEC in the AABC consortium.

**5.9 Women’s Circle of Health Study (WCHS)**

The WCHS has been described above. After quality control, we included 63 cases and 21 controls of African ancestry from WCHS in the AABC consortium.

**6. Ghana Breast Health Study (GBHS)**

The GBHS has been described above. The GBHS samples were genotyped using Infinium Global Screening Array-24. After quality control, we included 660 cases and 1,496 controls of African ancestry from GBHS genotyping data.

**7. Genetic Associations and Mechanisms in Oncology (GAME-ON) Consortium**

The GAME-ON Consortium consists of samples from NBHS, CBCS, NC-BCFR, MEC, PLCO, 2SISTER, SISTER, USRT, and WAABCS. The samples in GAME-ON Consortium were genotyped using the Infinium OncoArray-500k BeadChip.

**7.1 The Sister Study (SISTER)**

The Sister Study is a prospective cohort study designed to address genetic and environmental risk factors for breast cancer by the National Institute of Environmental Health Sciences. From 2003 through 2009, 50,884 U.S. women, including and Puerto Ricans, were recruited through a national multimedia campaign and network of recruitment volunteers, breast cancer professionals, and advocates. Participants were women aged 35 to 74 years and had a sister diagnosed with breast cancer^28^. At enrollment, participants completed baseline questionnaires on medical and family history, lifestyle factors, and demographics. Blood samples were collected during a home visit by trained phlebotomists and shipped overnight to the Sister Study laboratory where they were processed to obtain serum and stored at −80°C. After quality control, we included 130 cases and 163 controls of African ancestry from SISTER in the OncoArray consortium.

**7.2 The Two Sister Study (2SISTER)**

The Two Sister Study is a family-based retrospective study developed from the Sister Study. The Two Sister Study recruited the case sisters in the Sister Study who were diagnosed within 4 years and had been younger than age 50 years at diagnosis^29^. After quality control, we included 42 cases of African ancestry from 2SISTER in the OncoArray consortium.

**7.3 The United States Radiologic Technologists (USRT) cohort**

The USRT is a cohort study for cancer incidence and mortality which recruited approximately 140,00 U.S. radiologic technologists who were certified for at least two years between 1926 and 1982^30^. Breast cancer cases were confirmed based on pathology or medical records. After quality control, we included 26 cases and 38 controls of African ancestry from USRT in the OncoArray consortium.

**7.4 Women of African Ancestry Breast Cancer Study (WAABCS)**

The WAABCS is a hospital-based case-control study originally started in Nigeria in 1998 and was expanded to Uganda and Cameroon in 2011 with the same questionnaires and protocol^31,32^. After quality control, we included 308 cases and 292 controls of African ancestry from WAABCS in the OncoArray consortium.

**7.5 Nashville Breast Health Study (NBHS)**

The NBHS has been described above. After quality control, we included 51 cases and 53 controls of African ancestry from NBHS in the OncoArray consortium.

**7.6 Carolina Breast Cancer Study (CBCS)**

The CBCS has been described above. After quality control, we included 614 cases and 570 controls of African ancestry from CBCS in the OncoArray consortium.

**7.7 Northern California Breast Cancer Family Registry (NC-BCFR)**

The NC-BCFR has been described above. After quality control, we included 69 cases of African ancestry from NC-BCFR in the OncoArray consortium

**7.8 Multiethnic Cohort Study (MEC)**

The MEC has been described above. After quality control, we included 605 cases and 607 controls of African ancestry from MEC in the OncoArray consortium.

**7.9 The Prostate, Lung, Colorectal, and Ovarian Cancer Screening Trial (PLCO)**

The PLCO has been described above. After quality control, we included 24 cases and 68 controls of African ancestry from PLCO in the OncoArray consortium.

**8. The Vanderbilt Biobank (BioVU)**

The DNA biobank at Vanderbilt University consists of DNA extracted from blood collected during routine clinical testing and linked de-identified medical records^33,34^. Samples from more than 90,000 individuals were genotyped using the Illumina MEGA-Ex chip. Breast cancer cases were identified from the electronic medical record systems. In this study, we only kept adult participants of African ancestry. After quality control, we included 118 cases and 2,600 controls of African ancestry from BioVU. In addition, 356 controls of African ancestry from BioVU were selected and frequently matched on with cases from BEST study.

**9. Black Women: Etiology and Survival of Triple-negative Breast Cancers (BEST) Study**

The BEST is a case-only study which recruited African-ancestry women who were diagnosed with invasive breast cancer at age ≤50 years between 2009 and 2012 and lived in Florida at the time of their diagnosis^35^. Breast cancer cases were identified through the Florida Cancer Registry. Participants provided a saliva sample through mail for DNA extraction and *BRCA* testing.

The BEST samples were genotyped using the Infinium OncoArray-500k BeadChip. Controls from BioVU were selected to match cases from BEST by age. Given that different genotyping arrays were used for BEST and BioVU samples, we only kept genotyped variants shared by both arrays. Other criteria of quality control were the same as MEGA genotyping samples. After quality control, we included 359 cases from BEST and 356 controls matched from BioVU.

**10. Data from Collaborative Oncological Gene-environment Study (iCOGS)**

NBHS and SCCS (have been described above) contributed some samples in the iCOGS. Samples were genotyped using the Illumina iSelect Genotyping Array. After quality control and excluding the overlapped samples, we included 19 cases and 42 controls from NBHS, 44 cases and 251 controls from SCCS in iCOGS.

#### **II. Description of Studies included in the Asia Breast Cancer Consortium (ABCC)**

1.1 **Shanghai Breast Cancer Genetics Study (SBCGS)**

The Chinese participants were drawn from Shanghai Breast Cancer Genetics Study (SBCGS), which consists of the Shanghai Breast Cancer Study (SBCS), Shanghai Breast Cancer Survival Study (SBCSS), Shanghai Endometrial Cancer Study (SECS, contributed control data only), and the Shanghai Women’s Health Study (SWHS), four large population-based studies in urban Shanghai. All participants provided written informed consent prior to interview, and institutional review boards of all institutes in both China and the United States approved the study.

**1.1.1 Shanghai Breast Cancer Study (SBCS)**

The SBCS is a two-phase (SBCS-I and SBCS-II) population-based case-control study that recruited incident patients with breast cancer and controls in urban Shanghai, China.^36,37^ The first phase (SBCS-I) recruited 1,602 eligible breast cancer cases and 1,724 eligible controls, from August 1996 to March 1998. Cases were recruited by a rapid case-ascertainment system and the population-based Shanghai Cancer Registry, and controls were randomly selected from the general population using the Shanghai Resident Registry. There were 1,459 cases (91.1%) and 1,556 controls (90.3%) who completed in-person interviews. Blood samples (10 ml from each woman) were obtained who completed the in-person interview (1,193 (82%) cases and 1,310 (84%) controls). A sample of exfoliated buccal cells was obtained using cotton swabs from virtually all study participants who did not provide a blood sample. The second phase (SBCS-II) recruited subjects between April 2002 and February 2005 using a protocol similar to the one used in the initial phase. Similar to the SBCS-I subjects, the majority of newly-recruited cases (n=1,932, 97.1%) and controls (n=1,857, 93.4%) provided a blood sample or an exfoliated buccal cell sample to the study. The modified mouthwash method initially reported by Lum A *et al*. was used.^38^ Eligibility criteria for study participation were identical for SBCS-I and SBCS-II except age. The age ranged from 25 to 65 years for SBCS-I, and from 25 to 70 years in SBCS-II.

**1.1.2 Shanghai Breast Cancer Survival Study (SBCSS)**

The SBCSS included 6,303 breast cancer cases ascertained via the population-based Shanghai Cancer Registry between April 2002 and December 2006.^36^ Information on known breast cancer risk factors as well as anthropometrics was collected by in-person interviews using a protocol and questionnaire similar to that used in the SBCS. Buccal cell samples were collected from 96% of study participants using the modified mouthwash method. There were 1,469 breast cancer patients participated in both SBCS-II and SBCSS due to the time overlap in the participant recruitment period.

**1.1.3 Shanghai Endometrial Cancer Study (SECS)**

The SECS is a population-based, case-control study of endometrial cancer conducted between January 1997 and December 2003 using a protocol similar to the SBCS, and only the community controls from the SECS were included in the present study.^36^ Eligible cases were identified through the population-based Shanghai Cancer Registry and controls were randomly selected from the general population of Shanghai using the Shanghai Resident Registry and were age frequency matched to cases. Detailed information was collected by in-person interviews and anthropometrics measurements were taken. A total of 1,039 controls provided a blood sample or buccal cell sample using the mouthwash method, and these women were included in SBCGS.

**1.1.4 Shanghai Women’s Health Study (SWHS)**

The SWHS is a population-based cohort study which recruited approximately 75,000 adult women from urban Shanghai between 1997 and 2000.^39^ A total of 56,831 subjects, 75.8% of those who completed baseline survey through an in-person interview, donated a blood sample. An exfoliated buccal cell sample was collected from an additional 8,934 (49.3%) of the 18,111 subjects who did not provide a blood sample at baseline. Genomic DNA was available for about 88% of cohort members. Cancer cases were identified via record linkage with the population-based cancer registry and data collected at the Vital Statistic Unit, followed by home visits or telephone calls if necessary to confirm the diagnoses. Cancer diagnoses were verified by a review of medical records obtained from the diagnosing hospital.

Participants in SBCGS have been genotyped by multiple arrays, including Affymetrix Genome-Wide Human SNP Array 6.0 (Affy6), the Asian ExomeChip, the Multi-Ethnic Global Array (MEGA), Illumina Infinium OncoArray-500K BeadChip (OncoArray), and Illumina iSelect Genotyping Array (iCOGS). Similar genotyping and QC procedures have been described previously.^36,40^ After imputation with the 1000 Genomes Project Phase 3 and QC exclusions, the final SBCGS dataset included 2,511 cases and 2,127 controls by Affy6, 1,458 cases and 2,866 controls by ExomeChip, 1,794 cases and 2,059 controls by MEGA, 767 cases and 923 controls by OncoArray, and 515 cases and 890 controls by iCOGS.

1.2 **Hwasun Cancer Epidemiology Study-Breast (HCES-Br)**

The Hwasun Cancer Epidemiology Study (HCES-Br) is a hospital-based case-control study to identify factors of the cancer development and clinical progression in a Korean population.^41,42^ The study included 3,387 female breast cancer cases diagnosed between April 2004 and February 2013 at Chonnam National University Hwasun Hospital, a cancer specified hospital in Jeollanam-do province, South Korea. Patients with secondary or recurrent tumor were excluded. Controls were 3,186 women who were randomly selected from among women with no previous cancer diagnosis at enrollment in the Namwon Study and the Dong-gu study, ongoing community-based cohort studies in South Korea.^43^ Genomic DNA was extracted from their peripheral blood. Demographics data and conventional factors of breast cancer were collected by structured questionnaire and review of medical records. All cases and control subjects provided the informed consent to participate in the study and Institutional Review Board of Chonnam National University Hwasun Hospital approved this study.

In the HCES-Br, there were 274 cases and 273 controls genotyped by MEGA and imputed with the 1000 Genomes Project Phase 3 data as reference.

**1.3** **Korea Precision Oncology Program (KPOP) - Breast Cancer**

The KPOP – Breast Cancer study is a study to investigate genetic mutation/variants distribution of hereditary breast/ovarian cancer and risk stratification for women with or without family history of breast cancer. In addition, the risk factors of breast cancer were studied in women, stratified by family history of breast cancer. All cases had a histologically confirmed diagnosis of invasive breast cancer or ductal carcinoma in situ. The breast cancer cases were recruited from breast cancer center and genetic counseling clinic, National Cancer Center in Korea between 2013 and 2018. The controls were recruited from health screening examinees from National Cancer Center between 2013 and 2016 and they were women free of any cancer. After obtaining informed consent, cases and controls were asked to complete questionnaire on reproductive factors, lifestyle factors, and family history of cancer and provided blood samples. After separating plasma, serum, and whole blood, samples were stored at -70°C until assayed. Overall, 1904 breast cancer cases and 1195 controls were recruited

The current study includes 963 cases and 921 controls from KPOP, which were genotyped by MEGA and imputed with the 1000 Genomes Project Phase 3 data as reference.

**1.4 The Biobank Japan Project (BBJ2)**

The BioBank Japan Project recruited around 200,000 patients with 47 diseases in Japan and collaboratively collected DNA and serum samples (https://biobankjp.org/english/index.html).^44,45^ There were a total of 5,552 breast cancer patients and 89,731 female controls registered in Biobank Japan. Control samples were from population-based prospective cohorts and samples without related diagnoses. Samples were genotyped using the Illumina HumanOmniExpressExome BeadChip or a combination of the Illumina HumanOmniExpress and HumanExome BeadChips, and imputed with the 1000 Genomes Project Phase 3 data as reference.^46^

There were a total of 5,552 breast cancer patients and 89,731 female controls in Biobank Japan.

**1.5 Seoul Breast Cancer Study (SeBCS)**

The SeBCS is a hospital-based case-control study conducted in two teaching hospitals in Seoul.^47,48^ Between 2001 and 2007, there were 2,342 patients with primary breast cancer recruited in the study. Information on known breast cancer risk factors and anthropometrics were collected by in-person interviews using a protocol and questionnaire. Medical charts were reviewed to verify clinical information. Eligible controls were derived from a large urban cohort included in the Korea Genome Epidemiology Study (KoGES), which was an ongoing cohort study that has sought to understand the causes and risk factors of disease in South Korea. A total of 2,052 controls were recruited between May 2006 and December 2007. They were frequency-matched to cases on the case’s age at diagnosis in five-year intervals. Using a structured questionnaire and a protocol similar to the SeBCS, trained interviewers collected the demographic characteristics of the controls, their family histories with regard to breast cancer in first-degree relatives, reproductive and menstrual factors, and life-style habits.

After QC, there were 2,165 cases and 2,052 controls genotyped by Affymetrix 6.0^49^, 1,103 cases and 1,107 controls genotyped by OncoArray from SeBCS.

**1.6 Hospital-based Epidemiologic Research Program at Aichi Cancer Center (HERPACC)**

The participants were recruited from a hospital-based case-control study conducted in Aichi, Japan.^54^ All incident breast cancer cases were newly diagnosed within 1 year from the first visit to the Aichi Cancer Center between 2001 and 2013. Controls were selected from pool of non-cancer patients who firstly visited Aichi Cancer Center between 2001 and 2011. Subjects with previous cancer history were excluded.

After QC, there were 282 cases and 283 controls genotyped by OncoArray, 694 cases and 1,376 controls genotyped by iCOGS from HERPACC.

**1.7 Korean Hereditary Breast Cancer (KOHBRA)**

The KOHBRA study is an ongoing cohort study since 2007 to examine high risk groups for hereditary breast cancer such as female breast cancer patients with a family history, ovarian cancer, or other coincidental cancers, male breast cancer patients, and family members of breast cancer patients with *BRCA1/2* mutation. Final dataset included selected 1,397 female cancer patients without *BRCA1/2* mutation among KOHBRA subjects recruited in 2007-2009.^58^

After QC, there were 1,464 cases and 665 controls genotyped by OncoArray from KOHBRA.

**1.8 Nagano Breast Cancer Study (NGOBCS)**

The Nagano Breast Cancer Study is a multicenter, hospital-based case-control study which was conducted from May 2001 to September 2005 at four hospitals in Nagano Prefecture, Japan.^64,65^ Cases were admitted to the four hospitals during the survey period, and were a consecutive series of women aged 20-74 years with newly diagnosed, histologically confirmed invasive breast cancer. Among the 412 eligible patients, 405 (98%) agreed to participate. Controls were selected from medical checkup examinees in two of the hospitals who were confirmed having no cancer, with one control matched for each case by age (within three years) and residential area during the study period. Only one declined to participate among potential control subjects. Written informed consent was obtained from 405 matched pairs. Since two controls refused to provide blood samples, the analysis was restricted to 403 matched pairs. Participants completed a self-administered questionnaire, which included questions on demographic characteristics, anthropometric factors, smoking habits, family history of cancer, physical activity, medical history, and menstrual and reproductive history. Dietary habits were investigated using a 136- item semi-quantitative food-frequency questionnaire, which was developed and validated in the Japanese population. The ER status of the patient’s breast cancer tissue was obtained from medical records. Hormone receptor positivity values were determined either as specified by the laboratory that performed the assay, in accordance with the laboratory’s written interpretation thereof, or both. The study protocol was approved by the Institutional Review Board of the National Cancer Center (Tokyo, Japan).

After QC, there were 369 cases and 366 controls genotyped by OncoArray from KOHBRA.

**1.9 Taiwanese Breast Cancer Study (TWBCS)**

The study is a part of an ongoing collaborative study with a focus on understanding the cause of breast cancer among Taiwanese.^68,69^ Breast cancer patients were recruited from those who were diagnosed and treated at the Tri-Service General Hospital or the Changhua Christian Hospital between March 2002 and August 2005. The controls were randomly selected from women who attended the same hospitals for a comprehensive health examination during the same period. If any evidence of breast cancer, precancerous lesions of breast or other cancers was found, the subject was excluded from the control group. Epidemiologic data were collected from the participants via a structured questionnaire by research nurses. Blood biospecimen was also collected. All the participants provided their informed consent before the data and sample collection.

After QC, there were 551 cases and 256 controls genotyped by OncoArray, 889 cases and 236 controls genotyped by iCOGS from TWBCS.

#### **III. Description of BCAC European-ancestry participants**

Summary statistics data of European descendants from studies involved in the BCAC OncoArray, iCOGS, and GWAS projects were obtained and utilized in the cross-ancestry meta-analysis. Among 82 studies from the BCAC, the OncoArray dataset included 80,125 female cases with breast cancer and 58,383 female controls of European ancestry, and the Collaborative Oncological Gene-environment Study (iCOGS) included 38,349 breast cancer cases and 37,818 controls.^70^ In addition, summary statistics from 11 other breast cancer genome-wide association studies were also used in the meta-analysis with a combined sample of 14,910 cases and 17,588 controls. The genotyping data were imputed by IMPUTE version 2^71^ with the 1000 Genomes Project Phase 3 as the reference panel.

#### **IV. siRNA knockdown experiments**

The cell lines MDA-MB-231 and MCF-7 were obtained from the American Type Culture Collection (ATCC), which the cells were authenticated by short tandem repeat. The T47D cell line was a kind gift from Dr. Jennifer Pietenpol from Vanderbilt University Medical Center and was authenticated by short tandem repeat assay by ATCC.

The siRNA SMARTpools for each gene were obtained from the Vanderbilt High-Throughput Screening Facility’s Dharmacon ON-TARGETplus whole-genome siRNA library and reverse-transfected into each cell line using DharmaFECT 1 transfection reagent into a 384-well plate. Non-targeted (NT) siRNAs and siDEATH siRNAs (Qiagen) were used as negative and positive controls, respectively. Cells were incubated at 37 degrees for three days, nuclear stained with Hoechst dye for viable cell counts and propidium iodide for marking dead cells. Whole images of wells were captured using the Molecular Devices’ ImageXpress Micro XLS system using 4x objective lens and DAPI channel filter for nuclear stain and TexasRed filter for PI (dead cells). Subsequent cell viability was performed for each well (gene) by counting viable cell nuclei (+DAPI/-PI), using the MetaXpress software.

### **Acknowledgments**

The content is solely the responsibility of the authors and it does not necessarily represent the official views of the funding agents, including the U.S. Department of Health and Human Services, the National Institutes of Health, the National Cancer Institute, or the state cancer registries. The mention of trade names, commercial products, or organizations does not imply endorsement by the U.S. Government. The funders had no role in study design, data collection and analysis, decision to publish, or preparation of the manuscript.

The siRNA knock-down experiment was supported in part by the US National Institutes of Health grants R50CA211206 and S10OD028719 (JAB). Provided below are research supports for participating studies of the African-ancestry Breast Cancer Genetic (AABCG) Consortium and Asia Breast Cancer Consortium (ABCC).

**African-ancestry Breast Cancer Genetic (AABCG) Consortium**

The SCCS was supported by grant U01 CA202979 from the National Institute of Health (NIH). The NBHS was supported by NIH grant R01CA100374. The STSBHS was supported by NIH grants U54CA163069, U54CA163072 and R03CA192214 and the National Center for Advancing Translational Sciences (UL1TR000445). The BEST was supported by the National Cancer Institute (R01CA202981) and the Susan G. Komen Foundation (SAC210105). Vanderbilt University Medical Center’s BioVU projects were supported by numerous sources: institutional funding, private agencies, and federal grants. These include NIH funded Shared Instrumentation Grant S10OD017985, S10RR025141, and S10OD025092; CTSA grants UL1TR002243, UL1TR000445, and UL1RR024975. Genomic data are also supported by investigator-led projects that include U01HG004798, R01NS032830, RC2GM092618, P50GM115305, U01HG006378, U19HL065962, R01HD074711. The BWHS was supported by NIH grants U01CA164974, NIH R01CA228357, Susan G. Komen SAC 220228 (JRP), Karin Grunebaum Cancer Research Foundation (JRP). The authors would like to acknowledge contributions from central cancer registries supported through the Centers for Disease Control and Prevention’s National Program of Cancer Registries (NPCR) and/or the National Cancer Institute’s Surveillance, Epidemiology, and End Results (SEER) Program. Central registries may also be supported by state agencies, universities, and cancer centers. Participating central cancer registries include the following: AL, AR, AZ, CA, CO, CT, DE, DC, FL, GA, HI, IA, IL, IN, KY, LA, MD, MA, MI, MO, MS, NE, NJ, NM, NY, NC, OH, OK, OR, PA, SC, TN, TX, VA, WA, WI. The WCHS was supported by R01 CA10059, P01 CA151135, R01 CA185623 and the Breast Cancer Research Foundation. The MDABCS was supported in part from the Texas Tobacco Settlement Funds and the University of Texas MD Anderson Cancer Center Duncan Family Institute for Cancer Prevention and Risk Assessment. NBCS, WAABCS, and CCPS investigators were supported by National Institutes of Health (R01-CA89085, R01-CA142996, R01-CA228198, R01-CA242929, P20-CA233307, and R01-MD013452) and Breast Cancer Research Foundation (BCRF-22-071). The CBCS was supported by the University Cancer Research Fund of North Carolina, the Susan G Komen Foundation, the National Cancer Institute of the National Institutes of Health (P01CA151135), and the National Cancer Institute Specialized Program of Research Excellence (SPORE) in Breast Cancer (NIH/NCI P50-CA058223). The Northern California site of the Breast Cancer Family Registry (NC-BCFR) was funded by grant U01 CA164920 from the National Cancer Institute. The content of this manuscript does not necessarily reflect the views or policies of the National Cancer Institute or any of the collaborating centers in the Breast Cancer Family Registry (BCFR), nor does mention of trade names, commercial products, or organizations imply endorsement by the U.S. Government or the BCFR. The San Francisco Bay Area Breast Cancer Study (SFBCS) was supported by grants R01 CA63446 (E.M. John) and R01 CA77305 (E.M. John) from the National Cancer Institute, grant DAMD17-96-1-6071 (E.M. John) from the U.S. Department of Defense and grant 7PB-0068 (E.M. John) from the California Breast Cancer Research Program. The Women's CARE Study was supported in part by grants from the Breast Cancer Research Foundation (BCRF-24-132), Tower Cancer Research Foundation (Jessica M. Berman Senior Investigator Award), a gift from Dr. Richard Balch, and an endowed chair, the Harold E. Lee Chair for Cancer Research, and the USC Norris Comprehensive Cancer Center (NCI Cancer Center Support Grant P30 CA014089). The SISTER study was supported by the Intramural Research Program of the National Institutes of Health, National Institute of Environmental Health Sciences (grants Z01-ES044005 to D.P.S. and Z01-ES102245 to C.R.W.), Susan G. Komen for the Cure (grant FAS0703856 to C.R.W.). The BBCS was supported by NCI Center for Cancer Research Intramural Research Program (ZIA BC 010887). The NYUWHS was supported by NIH grant U01CA182934.

The GBHS project was funded with intramural funds from the National Cancer Institute, National Institutes of Health. GBHS would like to acknowledge the contribution from the following individuals to the GBHS: Korle Bu Teaching Hospital,Accra—Prof. Joe Nat Clegg-Lamptey, Dr. Florence Dedey, Dr. Lawrence Edusei, Dr. Verna Vanderpuye, Dr. Joel Yarney, Dr Adu-Aryee, Obed Ekpedzor, Angela Kenu, Victoria Okyne, Naomi Oyoe Ohene Oti, Evelyn Tay; Komfo Anoyke Teaching Hospital, Kumasi—Dr. Ernest Adjei, Dr. Francis Aitpillah, Dr. Daniel Ansong, Dr. Baffour Awuah, Dr. Joseph Oppong, Dr. Ernest Osei-Bonsu, Dr. Nicholas Titiloye, Marion Alcpaloo, Bernard Arhin, Emmanuel Asiamah, Isaac Boakye, Samuel Ka-chungu and; Peace and Love Hospital, Kumasi—Dr. Beatrice Addai Wiafe, Dr. Seth Wiafe, Samuel Amanama, Emma Abaidoo, Prince Agyapong, Thomas Agyei, Debora Boateng-Ansong, Margaret Frempong, Bridget Nortey Mensah, Richard Opoku, and Koﬁ Owusu Gyimah; University of Ghana, Accra—Prof. Richard Biritwum, Dr. Kofi Nyarko; and Dr. Jonine Figueroa of the University of Edinburg, Scotland. The study was further enhanced by the surgical expertise provided by Dr Lisa Newman of the University of Michigan and by pathological expertise provided by Drs. Stephen Hewitt and Petra Lenz of the National Cancer Institute and Dr. Maire A. Duggan from the Cumming School of Medicine, University of Calgary, Canada. Study management assistance was received from Ricardo Diaz, Shelley Niwa, Usha Singh, Ann Truelove and Michelle Brotzman at Westat, Inc. Appreciation is also expressed to the many women who agreed to participate in the study and to provide information and biospecimens in hopes of preventing and improving outcomes of breast cancer in Ghana. The GBHS also acknowledges the research contributions of the Cancer Genomics Research Laboratory for their expertise, execution, and support of this research in the areas of project planning, wet laboratory processing of specimens, and bioinformatics analysis of generated data. This project has been funded in whole or in part with Federal funds from the National Cancer Institute, National Institutes of Health, under NCI Contract No. 75N910D00024.

**Asia Breast Cancer Consortium (ABCC)**

Studies conducted among Asian women include (Principal Investigator, grant support): the Shanghai Breast Cancer Study (W.Z. and X.-O.S., R01CA064277), the Shanghai Women’s Health Study (W.Z., R37CA070867 and UM1CA182910), the Shanghai Breast Cancer Survival Study (X.-O. S., R01CA118229), the Shanghai Endometrial Cancer Study (X.-O.S., R01CA092585, controls only), the Seoul Breast Cancer Study [D.K., BRL (Basic Research Laboratory) program through the National Research Foundation of Korea funded by the Ministry of Education, Science and Technology (2012-0000347)], the BioBank Japan Project (S.-K.L., the Ministry of Education, Culture, Sports, Sciences and Technology from the Japanese Government); the Hwasun Cancer Epidemiology Study-Breast (S.-S.K., the Biobank of Chonnam National University Hwasun Hospital, a member of the Korea Biobank Network, # 07SA2014020); the KOHBRA/KOGES supported by a grant from the National R&D Program for Cancer Control, Ministry for Health, Welfare and Family Affairs, Republic of Korea (#10203) the Nagano Breast Cancer Study (M.I., National Cancer Center Research and Development Fund), the Hospital-based Epidemiologic Research Program at Aichi Cancer Center [Grant-in-Aid for Scientific Research on Priority Areas of Cancer (No. 17015018) from the Japanese Ministry of Education, Culture, Sports, Science and Technology and the “Practical Research for Innovative Cancer Control (15ck0106177h0001)” from the Japan Agency for Medical Research and development, AMED (K. Matsuo), and Cancer Bio Bank Aichi; and the Taiwanese Breast Cancer Study (C.-Y.S., the Taiwan Biobank project of the Institute of Biomedical Sciences, Academia Sinica, Taiwan).

Studies conducted among European-ancestry women Genotyping of the OncoArray was principally funded by three sources: the PERSPECTIVE project, funded from the Government of Canada through Genome Canada and the Canadian Institutes of Health Research, the Ministère de l’Économie, de la Science et de l’Innovation du Québec through Genome Québec, and the Quebec Breast Cancer Foundation; the NCI Genetic Associations and Mechanisms in Oncology (GAME-ON) initiative and Discovery, Biology and Risk of Inherited Variants in Breast Cancer (DRIVE) project [NIH Grants U19 CA148065, X01HG007492]; and Cancer Research UK [C1287/A10118, C1287/A16563]. The BCAC is funded by Cancer Research UK [C1287/A16563], the European Community’s Seventh Framework Programme under grant agreement 223175 [HEALTH-F2- 2009-223175] (COGS).
